# Post-processing and weighted combination of infectious disease nowcasts

**DOI:** 10.1101/2024.08.28.24312701

**Authors:** André Victor Ribeiro Amaral, Daniel Wolffram, Paula Moraga, Johannes Bracher

## Abstract

In infectious disease surveillance, incidence data are frequently subject to reporting delays and retrospective corrections, making it hard to assess current trends in real time. A variety of probabilistic nowcasting methods have been suggested to correct for the resulting biases. Building upon a recent comparison of eight of these methods in an application to COVID-19 hospitalization data from Germany, the objective of this paper is twofold. Firstly, we investigate how nowcasts from different models can be improved using statistical post-processing methods as employed, e.g., in weather forecasting. Secondly, we assess the potential of weighted ensemble nowcasts, i.e., weighted combinations of different probabilistic nowcasts. These are a natural extension of unweighted nowcast ensembles, which have previously been found to outperform most individual models. Both in post-processing and ensemble building, specific challenges arise from the fact that data are constantly revised, hindering the use of standard approaches. We find that post-processing can improve the individual performance of almost all considered models both in terms of evaluation scores and forecast interval coverage. Improving upon the performance of unweighted ensemble nowcasts via weighting schemes, on the other hand, poses a substantial challenge. Across an array of approaches, we find modest improvement in scores for some and decreased performance for most, with overall more favorable results for simple methods. In terms of forecast interval coverage, however, our methods lead to rather consistent improvements over the unweighted ensembles.

## 1 Introduction

Real-time surveillance plays a critical role in monitoring and analyzing the spread of infectious diseases, but the availability of timely and accurate data remains a challenge. The nature of data collection and reporting introduces delays, which cause recent data points to be incomplete and trends difficult to assess. Statistical nowcasting methods can be employed to predict by how much recent values will be corrected upwards.

Such methods have been extensively employed in various infectious disease settings, including dengue (Codeco et al., 2018; Bastos et al., 2019; Beesley et al., 2022), HIV (Cox and Medley, 1989) and outbreaks of gastrointestinal diseases (Höhle and an der Heiden, 2014). During the COVID-19 pandemic, the topic received increased attention (Greene et al., 2021; Günther et al., 2021; Seaman et al., 2022; Lison et al., 2024) as many countries and health authorities faced similar challenges. The present work builds upon a systematic comparison of nowcasting methods in a real-time application to German COVID-19 hospitalization incidences (Wolffram et al., 2023). For this study, a complete set of daily probabilistic nowcasts from eight models and over a six-month period (from November 2021 to April 2022) was compiled, which we use to study two related research questions.

Firstly, we develop statistical post-processing methods for infectious disease nowcasts, similar to existing methods from weather forecasting (Gneiting et al., 2005; Schulz et al., 2021). Post-processing aims at correcting systematic shortcomings of predictions from individual models, like biases and dispersion errors. In our case study, underdispersion of forecasts, i.e., too narrow prediction intervals, was the most common shortcoming of models. In order to suitably transform model outputs, an additional statistical model is fitted to past nowcast and observation pairs. Secondly, we address ensemble nowcasts, which combine different individual nowcasting models. Simple unweighted nowcast ensembles have been found to perform favourably in Wolffram et al. (2023), raising the question whether further improvements can be achieved by weighting different models in a suitable manner. Data-driven weighting of ensemble members is an active area of research in infectious disease forecasting (Yamana et al., 2016; Reich et al., 2019; Reis et al., 2019). For instance the US CDC have used weighted forecast ensembles to inform public health decision making during the COVID-19 pandemic (Ray et al., 2023). To date, however, evidence on the benefits relative to simple unweighted ensembles remains mixed (Bracher et al., 2021a; Ray et al., 2023). This echoes the broader statistical literature, where it has been pointed out that the estimation of ensemble weights comes at a cost which may not necessarily be outweighed by the benefits (Claeskens et al., 2016).

In our application to German COVID-19 hospitalization incidences, we find that post-processing of infectious disease nowcasts leads to quite consistent improvements across nowcasting methods and horizons. This holds both for nowcast calibration in terms of interval coverage rates and for score-based evaluation. Data-driven weighting of nowcast ensembles, on the other hand, proves to be a very challenging task. Exploring a variety of weighting methods, we find consistent improvements in calibration. In terms of evaluation scores, however, we obtain modest improvements for some approaches, and considerable deterioration of performance for others. The more successful weighting schemes tend to be simple, while added complexity rarely translates to improvements.

The remainder of this paper is structured as follows. In Section 2, we describe our applied setting and highlight the challenges of dealing with incomplete data. In Section 3, we introduce the notation used throughout the paper, present the post-processing and ensemble modeling approaches, and discuss the specific challenges posed by data revisions. Section 4 shows the obtained results based on the previously introduced post-processing and ensemble methods applied to the German COVID-19 hospitalization data. Lastly, in Section 5, we discuss our results and comment on the limitations and possible extensions of our work.

## 2 Motivation: COVID-19 hospitalizations in Germany

For illustration we briefly sketch our applied nowcasting setting, to which we will return in Section 4. We are concerned with the *7-day COVID-19 hospitalization incidence* (German Federal Ministry of Health, 2023). These data, updated daily by Robert Koch Institute (2022), played an important role in pandemic planning in Germany especially in fall and winter 2021/2022. Temporarily, this indicator even served to determine the necessary level of non-pharmaceutical interventions via a set of thresholds (German Federal Government, 2023). The 7-day hospitalization incidence is defined as the number of new COVID-19 cases registered by local health authorities over a 7-day period which ultimately led to a hospitalization. Hospital admission is not required to have taken place during the same 7-day period and may in fact occur considerably later. This somewhat unintuitive definition, which was chosen as “a compromise between timeliness and data quality” (Norddeutscher Rundfunk, 2023), implies that hospitalization counts are not aggregated by the day of admission, but by the day of case registration (see Section 2.1 of Wolffram et al. 2023 for a more detailed account). As a consequence, the delay problem described in Section 1 is particularly pronounced for this indicator: an additional delay between the date of case registration and the date of admission is added on top of the actual reporting delay for the hospitalization. This results in strongly incomplete values of the hospitalization incidence for recent dates, and a characteristic dip at the end of the time series. As detailed in Wolffram et al. (2023), data are corrected upwards over prolonged periods of time, and may still change months after initial reporting.

Figure 1 illustrates the nowcasting task and nowcasts generated in real time using three different methods. The black lines show data as available when the respective nowcast was issued. The red line shows a later version of the time series including retrospective completions. Light grey lines show unrevised data where for each date only the initial value reported on that same date is shown (implying that the latest value of the black and grey lines coincide). Nowcasts, i.e., predictions of completed incidence values, are shown as coloured bands. These have been collected in the *German COVID-19 Nowcast Hub* (https://covid19nowcasthub.de), a collaborative modelling project involving eight independent modelling groups. The Nowcast Hub aimed to provide reliable assessments of recent trends via daily updated nowcasts, but also to conduct a systematic methods comparison (Wolffram et al., 2023). The analyses in the present paper will be based on the study period of this comparative evaluation (November 29, 2021, through April 29, 2022). Overall, we consider eight different individual (i.e., stand-alone) models from the project, which are described briefly in Section SS1 (Supplementary Material). Moreover, unweighted median and mean ensembles are available, see Section 3.5.1.

**Figure 1:**
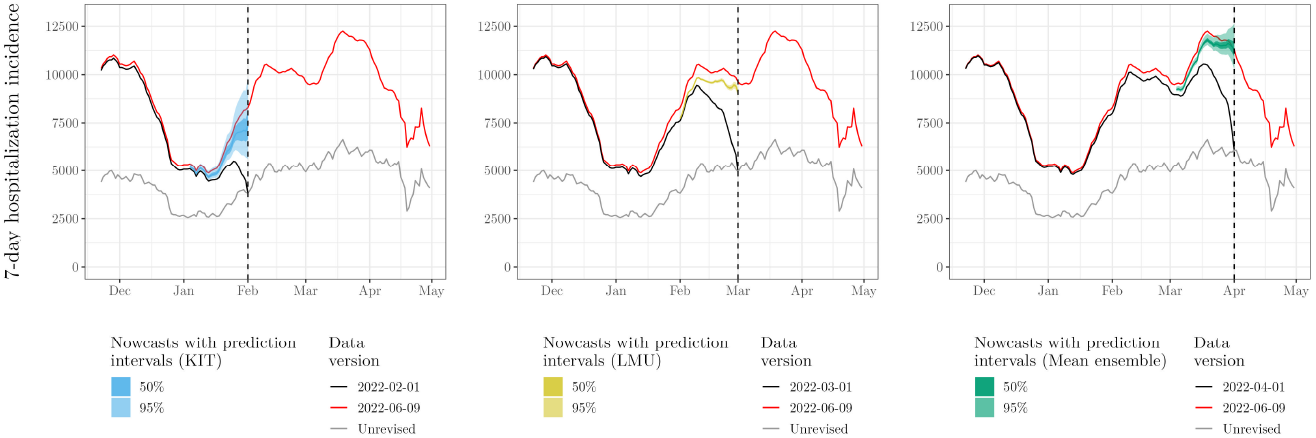
Illustration of the nowcasting task and nowcasts from three different models (`KIT`, `LMU`, and a mean ensemble) on February 01, March 01, and April 01, 2022, respectively. Black lines show data as available in real time on the respective forecast date, with the characteristic dip due to delays. The red line shows the data as completed later (40 days after the end of the displayed period). Point nowcasts and 50% and 95% uncertainty intervals are shown in colors.

As can be seen from Figure 1, different methods produce nowcasts with different characteristics. The `KIT` model, shown in the left panel, issued rather wide uncertainty intervals, while the intervals from the `LMU` model (middle panel) were considerably more narrow. The right panel shows the median ensemble nowcast, which represents an unweighted combination of all eight models and has uncertainty intervals of medium width.

## 3 Methods

In this section, we introduce basic concepts and notation on probabilistic disease nowcasts and their evaluation. Moreover, we describe the methods employed for post-processing and ensemble forecasting, and discuss the particularities arising from the fact that observations are subject to revisions.

### 3.1 Notation for probabilistic nowcasting

Denote by *x*_1_, …, *x*_*T*_, a daily time series of interest. In our application, *x*_*t*_ is a rolling sum over trailing 7-day windows, but is nonetheless indexed by days. We assume that *x*_*t*_ is not directly observable in real time. Instead, on day *t*, we observe a preliminary version 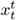. This value is subsequently revised each day, with 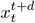 denoting the value as available on day *t* + *d*. We assume that data are only subject to revisions up to *D* days after the fact, so that

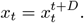

In our application, we use *D* = 40, and as revisions arise from delayed reports they are typically upwards. The hospitalizations added to the record with a delay of *d* days correspond to the increment 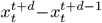. It is common to arrange the increments in a *reporting triangle* (Günther et al., 2021), but for our purposes it is more straightforward to use the above notation.

At time *t*^⋆^, the nowcasting task consists in predicting 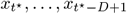, i.e., the final values of those data points which are still subject to revisions. Nowcasts are typically based on the corresponding partial data 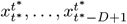, but may also take into account other information available at *t*^*^. Throughout the paper, we will consider probabilistic nowcasts, stored as quantiles at pre-defined levels *α*_1_, …, *α*_*A*_ (in our application, these are 0.025, 0.1, 0.25, 0.5, 0.75, 0.9, 0.975). For each level *α*, we denote the predictive *α* quantile for *x*_*t*_ issued by model *m* at time *t*^⋆^ by

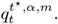

In the following, we refer to day *t*^*^ as the “nowcast date” and day *t* as the “target date.” Moreover, we denote by *h* = *t* − *t*^⋆^ the *horizon* of the nowcast, meaning that on day *t*^*^, a nowcast horizon of *h* = 0 days refers to target date *t*^*^, *h* = −1 day refers to the previous day *t*^*^ − 1 and so on. Consequently, for nowcast horizon *h* = 0, only the initial reports are known at the time of nowcasting, while for *h* = −1 reports with one day of delay are already available etc. Note that unlike in classical forecasting settings, the horizons are negative in nowcasting, and to enhance readability we will usually write “1 day back” rather than “horizon *h* = −1 day” etc.

### 3.2 Evaluation metrics

Post-processing and ensemble weighting typically require assessing the historical predictive performance of different models. To this end, we will employ the weighted interval score (WIS, Bracher et al. 2021a), which has been widely used to evaluate quantile-based predictions during the COVID-19 pandemic (e.g., Cramer et al. 2022). Denote by *F* a predictive distribution issued for a quantity *x*, and by 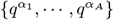 the available quantiles of *F*. The WIS is built upon the piece-wise linear quantile score (Gneiting and Raftery, 2007), also known as the “pinball loss.” For quantile level *α*, it is given by

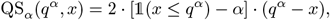

where 𝟙 denotes the indicator function. The WIS is defined as the average quantile score across levels,

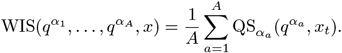

The WIS is negatively oriented, i.e., smaller values are better. It represents a quantile-based approximation of the continuous ranked probability score (CRPS; Gneiting and Raftery 2007) and can be interpreted as a probabilistic generalization of the absolute error. It is a proper scoring rule, meaning that it encourages honesty of forecasters. As detailed in Bracher et al. (2021a) and Section SS2 (Supplementary Material), the WIS can be split into components for forecast spread, overprediction, and underprediction. This will be used to characterize biases and dispersion errors of different models.

As in Wolffram et al. (2023), we use *relative WIS* values with respect to a naïve baseline model to put average scores into perspective. Here, the naïve baseline simply consists in setting all nowcast quantiles to the currently known incomplete data value (i.e., our baseline corresponds to simply ignoring reporting delays). The relative WIS is defined as

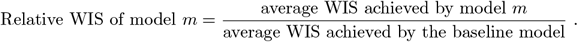

E.g., in meteorology it is common to report *skill scores*, which correspond to “1 − relative WIS”. We here prefer the relative WIS as it is easily displayed along with average scores using a second axis.

In addition to score-based evaluation, we assess the probabilistic calibration of nowcasts via interval coverage fractions (i.e. fraction of cases in which prediction intervals contained the true value). These are reported for the central 50% and 95% prediction intervals.

### 3.3 Including preliminary observations in nowcast evaluations

In nowcasting, information on the target quantity accumulates more gradually than in classical forecasting. On day *t*^*^, the WIS thus cannot be evaluated for target dates *t*^*^ − 1, …, *t*^*^ −*D*, even though some new information on 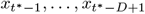 has already accumulated, with, e.g., 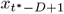 usually almost exactly known. Simply ignoring the respective nowcasts in performance assessment would mean giving up on information which due to its recency may be particularly relevant. We will assess the two following approaches to integrate it into our post-processing or ensemble weighting methods.

- **Simple imputation:** In order to complete the partial observations 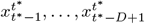 on day *t*^*^, an obvious strategy is to use up-to-date nowcasts. We thus employ pseudo-observations defined as

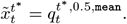 We use predictive medians from the unweighted mean ensemble, denoted by `mean`, which we know has rather reliable performance (Wolffram et al., 2023). Intuitively speaking, rather than comparing nowcasts issued during the last *D* − 1 days to the truth, we assess how strongly they already had to be revised in light of new data.
- **Imputation with uncertainty:** The simple imputation approach neglects the uncertainty remaining in the <monospace>mean</monospace> ensemble nowcasts. In a second, more sophisticated approach, we compare past nowcasts to all quantiles 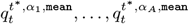. This can be done using a generalization of the WIS described in Section SS3 (Supplementary Material). It is inspired by a similar generalization of the CRPS which has been suggested by Friederichs and Thorarinsdottir (2012) to account for observation errors in meteorological forecast evaluation.

### 3.4 Post-processing individual models

We now address the improvement of nowcasts from individual models via statistical post-processing. To this end, we employ a simple re-scaling approach. Specifically, at nowcast time *t*^*^, the predictive *α* quantile issued by a given model for target time *t* is transformed as

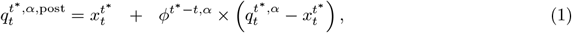

where we suppressed the index *m* for the model. Scaling is thus only applied to the difference between the currently known value 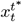 and the predicted 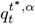. In our application, we will constrain *ϕ*^*h,α*^ *>* 0, which ensures that the nowcast quantile cannot fall below the already known number of hospitalizations. In the most general formulation, the scaling *ϕ*^*h,α*^ is specific to the quantile level *α* and the nowcast horizon *h*. While we also consider a more parsimonious formulation where a shared *ϕ*^*α*^ is used across horizons, we always keep it specific to *α*. The reason is that in case of of dispersion errors, corrections need to be upward for some quantile levels and downward for others.

The value of *ϕ*^*h,α*^ is determined via score minimization over a training period *R*, i.e., it is chosen such that the objective

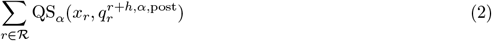

is minimized. The set ℛ includes days *t*^*^ − *R*, …, *t*^*^ − *D* for which definitive observations are available. In our main analysis we use *D* = 40 and *R* = 90 days if individual-model nowcasts have been available for this long. Otherwise we set *R* to the maximum feasible number, which we ensure to be at least 70. Depending on the strategy chosen to handle incomplete data, *ℛ* may in addition contain days *t*^*^ − *D* + 1, …, *t*^*^ − 1, for which pseudo-observations are employed in the evaluation. In the case of imputation with uncertainty, we use the previously mentioned adaptation of the quantile score from Section SS3 (Supplementary Material). As in Ray et al. (2023), we determine *ϕ*^*h,α*^ via a grid search.

### 3.5 Combination of nowcasting models

To combine nowcasts from *M* models into an ensemble we use mappings of the form

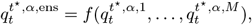

i.e., the ensemble quantile is computed from the respective member quantiles at the same level. In the following, we elaborate on different specifications of *f*, from simple unweighted to sophisticated data-driven schemes. As discussed e.g., in Ray et al. (2023), the space of possible formulations and parameterizations is vast. Our rationale is to explore a set of distinct, but reasonably simple approaches which could be operated in practice.

#### 3.5.1 Unweighted combination

The simplest approach is given by unweighted aggregation, as in the mean ensemble defined by

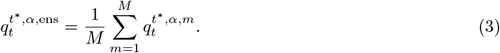

Paralleling Wolffram et al. (2023), we will also consider a median ensemble, which uses the median rather than the mean to aggregate quantiles from different models. We opt for direct aggregation of predictive quantiles, also called *Vincentization* (Genest, 1992), as the available submissions consist exclusively of quantiles. This makes it difficult to compute, e.g., linear pools or other forms of mixture distributions. Vincentization is commonly used in collaborative disease forecasting (see e.g., Ray et al. 2023), and more details on its properties can be found in Lichtendahl et al. (2013).

#### 3.5.2 Post-processing-based approaches

An obvious approach to improve upon the unweighted ensemble is to harness the post-processing methods described in Section 3.4. As the order of post-processing and combination of forecasts is not interchangeable, we consider two approaches:

- **Post-process, then combine:** If post-processing can improve upon individual models, one may expect a combination of post-processed models to be superior. We thus consider unweighted mean and median ensembles of the post-processed members.
- **Combine, then post-process:** Alternatively, the different models can be combined to an unweighted mean or median ensemble first, which is subsequently subject to post-processing. This is computationally cheaper as post-processing only needs to be run once.

#### 3.5.3 Direct inverse-score weighting

A second rather straightforward strategy consists in “direct inverse-score weighting” (DISW). We here generalize Equation (3) to

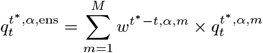

while choosing the weights in a heuristic manner, setting

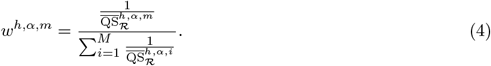

Here,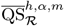 is the average quantile score for model *m*, quantile level *α* and horizon *h* days during the training period *ℛ* from Equation (2). The rationale is that models with good historical performance (low average scores) should receive larger weights. As in Section 3.4, we will also assess a version with weights *w*^*m,α*^ shared across horizons. Inverse-score weighting has been used for COVID-19 forecasts in Bracher et al. (2021b), where in turn it had been borrowed from the meteorological literature (Zamo et al., 2021). An advantage of this approach is that it does not require any costly optimization.

#### 3.5.4 Adjustable inverse-score weighting

Direct inverse score weighting has two obvious limitations. Firstly, it makes a strong assumption on how weights should depend on past WIS scores. Secondly, as it is a convex combination of the models, no correction for biases shared by all members is possible. If, for instance, all member models show a downward bias, then so will the ensemble. We therefore render the approach more flexible by introducing two additional parameters *ϕ*^*h,α*^ and *θ*^*h,α*^. We will refer to this as “adjustable inverse-score weighting” (AISW). Combining ideas from Equations (1) and (4), we set

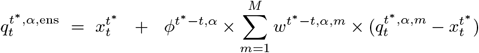

with weights defined as

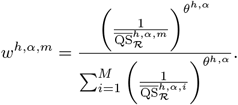

Here, 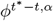 can shift predictive quantiles up and down. As in the post-processing scheme from Section 3.4, scaling is only applied to the predictions of yet-to-observe hospitalizations, while the current count 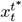 is not modified. If only one model is available, the approach is thus equivalent to Equation (1). The parameter *θ*^*h,α*^ steers how strongly weights depend on past performance. A value of 0 implies equal weighting as in Equation (3) (meaning that a simplified version of DISW with *θ*^*h,α*^ = 0 is the same as the post-processed mean ensemble from Section 3.5.2). Positive values of *θ*^*h,α*^ mean that more weight is given to models with good past performance. For *θ*^*h,α*^ = 1, the weights correspond to the DISW approach (4). Again, we also apply a simplified version where parameters are shared across horizons. The weights and scaling parameter are determined via score optimization and a grid search as in Equation (2).

This approach is a variation of the one from Ray et al. (2023). It keeps the number of parameters moderate and circumvents identifiability problems arising from strong correlations between quantiles from different models (indeed, unconstrained quantile regression was poorly behaved in our application). While Ray et al. (2023) use an exponential transformation 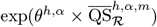, we opted for a power relationship 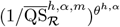. This way, Equation (4) nests into the general formulation. We compared the exponential and power formulations in exploratory analyses and found them to behave similarly.

#### 3.5.5 Top-*n* model selection

An alternative to explicit weighting is to restrict the ensemble to a pre-specified number *n* of models which have shown the best performance (or, put differently, to eliminate *M* − *n* models with weaker performance). At time *t*^*^ and for each quantile level *α* and horizon *h*, we thus order models according to the average quantile score 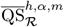. Then, the *n* best-performing models are retained and averaged into a mean or median ensemble without further weighting. We will explore different values of *n*, i.e., remove weaker models one by one. As for the other approaches, we will also consider a simplified version where all horizons are treated jointly.

## 4 Application to German COVID-19 hospitalizations

We now provide details on the COVID-19 hospitalization nowcasting task from Section 2 and highlight differences to previous work. This is followed by a performance assessment for the various proposed methods. To keep the presentation structured, we provide some interpretation of the results already in the respective subsections rather than the discussion part.

### 4.1 Technical description of the nowcasting task

#### Nowcasting horizons, stratification and target

Paralleling Wolffram et al. (2023), we will consider nowcasts up to 28 days back, i.e., at horizons *h* = 0, …, −28 days. These are available at the national level, for the 16 German states and for 7 age groups (0–4, 5–14, 15–34, 35–59, 60–79 and 80+ years; pre-defined by RKI). We consider delays up to *D* = 40 days, i.e., nowcasts for target date *t* aim to predict and are evaluated against

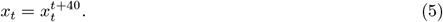

As mentioned in Section 3.2, nowcasts are stored as a set of quantiles at levels 0.025, 0.1, 0.25, 0.5, 0.75, 0.9, 0.975.

#### Study period

We consider nowcasts generated in a daily rhythm from November 29, 2021, to April 29, 2022. As all data-driven post-processing and ensembling methods require some historical pairs of nowcasts and observations for training, we hold out the first 70 days of this period. The performance evaluation is conducted over the remaining time period (February 8, 2022 through April 29, 2022; i.e., 81 days). By leaving out 70 days, we ensure that a minimum of 30 days of complete data is available for training the post-processing and ensembling methods.

#### Revision of nowcasting target definition

We note that in Wolffram et al. (2023), a different target definition was used, and we provide a brief justification for this change. The previous definition for target date *t* was the incidence value including all revisions made up to August 8, 2022 (i.e., 100 days after the last nowcasting date). With *t*_max_ as the index of August 8, 2022, this corresponds to

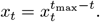

This was meant as a “final value”, based on the assumption that no further revisions would occur after this date. In reality, however, the data kept being revised upwards (Wolffram et al., 2023, Section 3.7). This is disadvantageous as revisions could accumulate over a longer time for target dates early in the study period (*t*_max_ − *t* = 181 days, for February 8, 2021) and were thus overall stronger than for later target dates (*t*_max_ − *t* = 100 days, for April 29, 2022). As already discussed in Wolffram et al. (2023), we therefore consider Equation (5) a more suitable and well-defined target. We opted for *D* = 40 as this was the maximum delay most modelling teams assumed in their statistical analysis. For the `ILM` team, who used *D* = 84, we obtained adjusted nowcasts with a matching maximum delay.

### 4.2 Performance of original nowcasts from Wolffram et al. (2023)

We start by briefly summarizing the performance of the eight individual models and two ensembles from Wolffram et al. (2023) in our adapted setting. Figure 2 shows nowcasts issued by different models over time for two horizons (0 and 14 days back). Figure 3 displays average WIS values and interval coverage fractions for national-level and stratified nowcasts. Note that the `ILM` and `RKI` teams did not report nowcasts for states and ages groups, respectively. This figure is similar to Figure 13 from Wolffram et al. (2023), but refers to our shortened evaluation period. For a more detailed account, we present results per age group along with comments for interpretation in Supplementary Section SS4.

**Figure 2:**
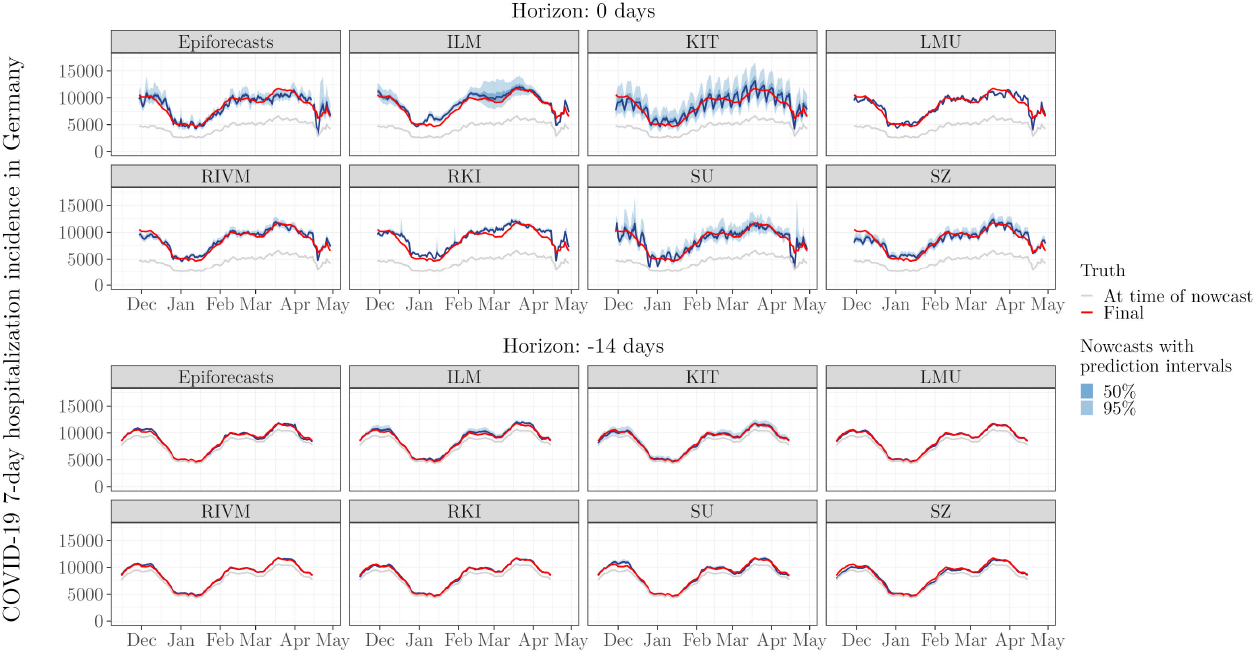
National-level nowcasts (blue) 0 and 14 days back for the eight individual models, by target date. The red line shows the nowcasting target, i.e., the number of COVID-19 7-day hospitalization cases after 40 days of retrospective corrections. The grey lines show the reported incidence counts at the time of nowcasting, i.e., after 0 (top) and 14 days (bottom), of retrospective corrections. Blue shaded areas represent nowcast intervals. This figure parallels Figures 5 and 6 from Wolffram et al. (2023).

**Figure 3:**
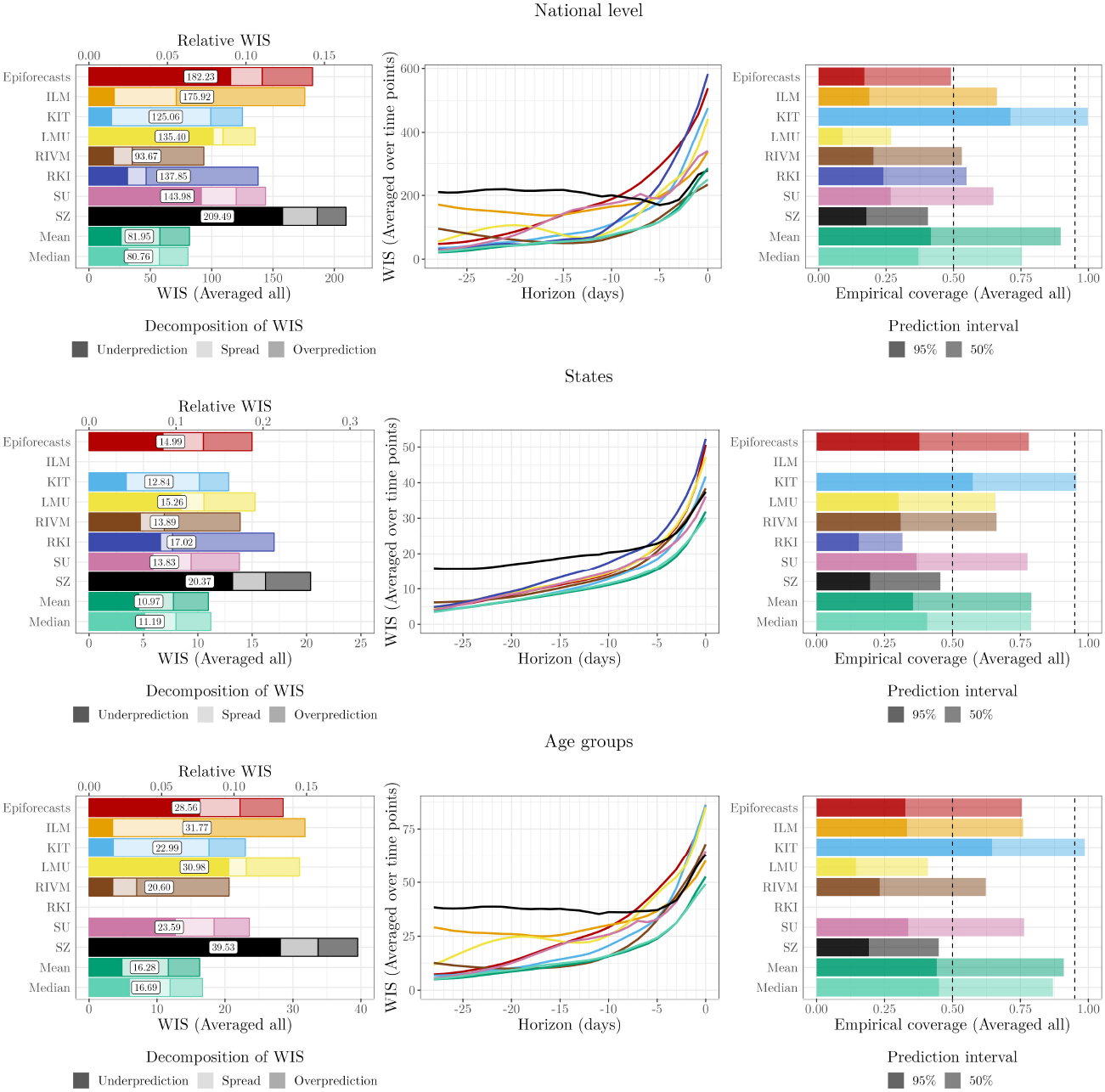
Model performance of original models and ensembles from Wolffram et al. (2023). Left: WIS (averaged over time points and horizons), split into components for underprediction, spread, and overprediction. A second axis at the top of the plot shows relative WIS with respect to a naïve baseline of no delay correction (see Section 3.2). Middle: WIS by nowcast horizon (averaged over time points). Right: Empirical coverage proportions (averaged over time points and horizons). The results are reported for the national level (top row) and averaged across states (middle row) and age groups (bottom row).

The mean and median ensembles achieve substantially better average WIS than all individual models. Also, their prediction intervals, while not reaching nominal coverage, are better calibrated. Most individual models have considerably too low interval coverage fractions (right column). This reflects overly narrow prediction intervals, as also indicated by the small dispersion components of the WIS. This pattern is particularly pronounced for the `LMU`, `RIVM` and `RKI` models, while the `KIT` model is somewhat better calibrated (see also Figure 2). The `SZ` model has a large underprediction component of the average WIS, suggesting a downward bias. We note that the WIS values for the stratified targets are lower on average because the WIS is scale-dependent.

### 4.3 Performance of post-processed individual models

We employed the methods from Section 3.4 to post-process the nowcasts from all eight individual models. In our main analysis, we used a maximum of *R* = 90 days for training. In the Supplementary Material, we present results for a maximum of *R* = 60 days and without any maximum value for *R* (finding that the improvements in average WIS when using data from more than *R* = 90 days are minor; see Figure SF2). Varying the analytical options described in Section 3.3, we investigated the post-processing approach with four different settings (see upper part of Table 1). These differ in how yet incomplete observations are included into the training set (Section 3.3) and whether the scaling parameters are shared across horizons. For each version, we introduce a label which we will use for referencing in the following (set in `typewriter` font).

**Table 1:**
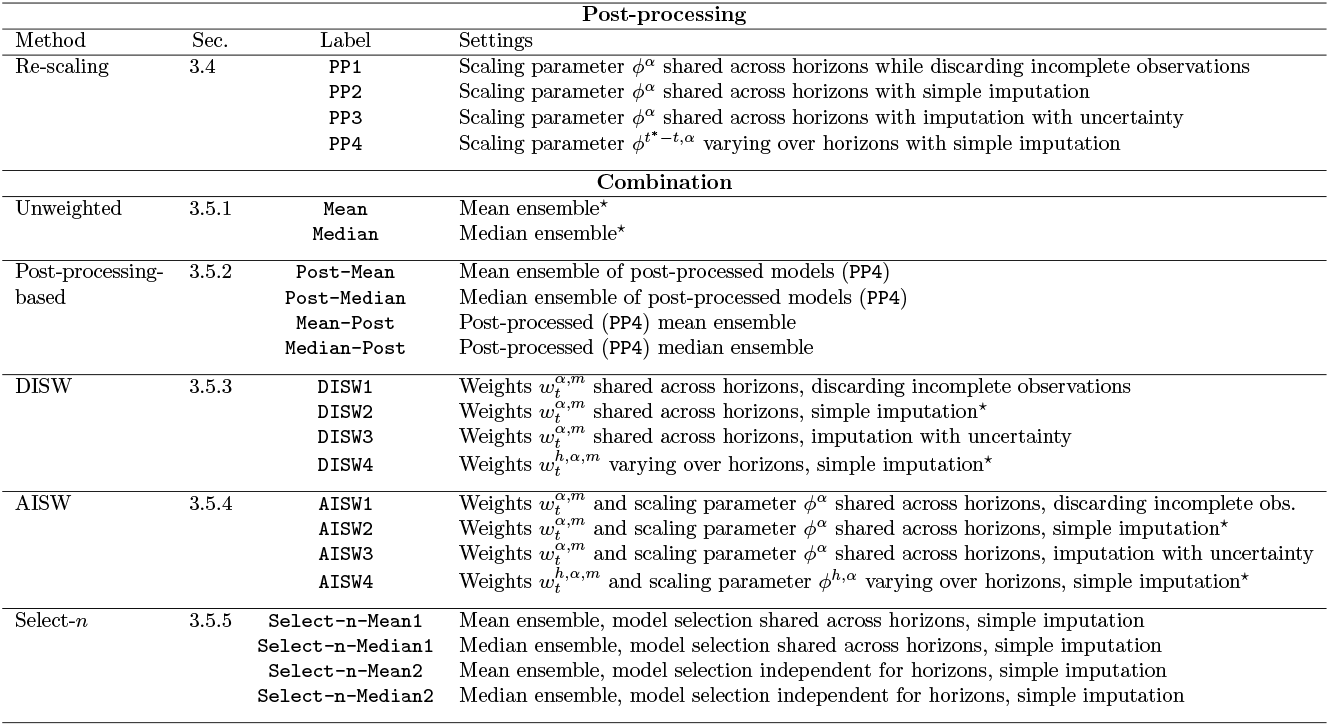
Post-processing and combination approaches assessed in Section 4. All methods are fitted to national-level data, methods marked with a star symbol (⋆) are moreover applied to stratified data (age groups and states). The “Label” column contains a short identifier used for brevity in the remaining text and figures.

| Post-processing |  |  |  |
| --- | --- | --- | --- |
| Method | Sec. | Label | Settings |
| Re-scaling | 3.4 | PP1 | Scaling parameter $\phi^\alpha$ shared across horizons while discarding incomplete observations |
| | | PP2 | Scaling parameter $\phi^\alpha$ shared across horizons with simple imputation |
| | | PP3 | Scaling parameter $\phi^\alpha$ shared across horizons with imputation with uncertainty |
| | | PP4 | Scaling parameter $\phi^{t^*-t,\alpha}$ varying over horizons with simple imputation |
| Combination |  |  |  |
| Unweighted | 3.5.1 | Mean | Mean ensemble* |
|  |  | Median | Median ensemble* |
| Post-processing-based | 3.5.2 | Post-Mean | Mean ensemble of post-processed models (PP4) |
|  |  | Post-Median | Median ensemble of post-processed models (PP4) |
|  |  | Mean-Post | Post-processed (PP4) mean ensemble |
|  |  | Median-Post | Post-processed (PP4) median ensemble |
| DISW | 3.5.3 | DISW1 | Weights $w_t^{\alpha,m}$ shared across horizons, discarding incomplete observations |
| | | DISW2 | Weights $w_t^{\alpha,m}$ shared across horizons, simple imputation* |
| | | DISW3 | Weights $w_t^{\alpha,m}$ shared across horizons, imputation with uncertainty |
| | | DISW4 | Weights $w_t^{h,\alpha,m}$ varying over horizons, simple imputation* |
| AISW | 3.5.4 | AISW1 | Weights $w_t^{\alpha,m}$ and scaling parameter $\phi^\alpha$ shared across horizons, discarding incomplete obs. |
| | | AISW2 | Weights $w_t^{\alpha,m}$ and scaling parameter $\phi^\alpha$ shared across horizons, simple imputation* |
| | | AISW3 | Weights $w_t^{\alpha,m}$ and scaling parameter $\phi^\alpha$ shared across horizons, imputation with uncertainty |
| | | AISW4 | Weights $w_t^{h,\alpha,m}$ and scaling parameter $\phi^{h,\alpha}$ varying over horizons, simple imputation* |
| Select-$n$ | 3.5.5 | Select-n-Mean1 | Mean ensemble, model selection shared across horizons, simple imputation |
|  |  | Select-n-Median1 | Median ensemble, model selection shared across horizons, simple imputation |
|  |  | Select-n-Mean2 | Mean ensemble, model selection independent for horizons, simple imputation |
|  |  | Select-n-Median2 | Median ensemble, model selection independent for horizons, simple imputation |

The average WIS and coverage proportions for the post-processed models are presented in Figure 4 for `PP4` and Supplementary Figures SF3–SF5 for the other settings. Quite consistently across post-processing specifications and models, the average WIS values decrease, the WIS components are more balanced and the coverage rates are closer to the nominal values. Comparing Supplementary Figures SF3 (`PP1`) and SF4 (`PP2`), we see that including yet incomplete observations into the training set is beneficial, yielding improved WIS performance for almost all models. The more sophisticated imputation with uncertainty (`PP3`, Figure SF5) considerably increased computation times, but compared to simple imputation (`PP2`) had limited impact on the nowcasts and their performance. The more flexible version `PP4` with separate handling of different horizons (Figure 4) results in slightly better overall performance.

**Figure 4:**
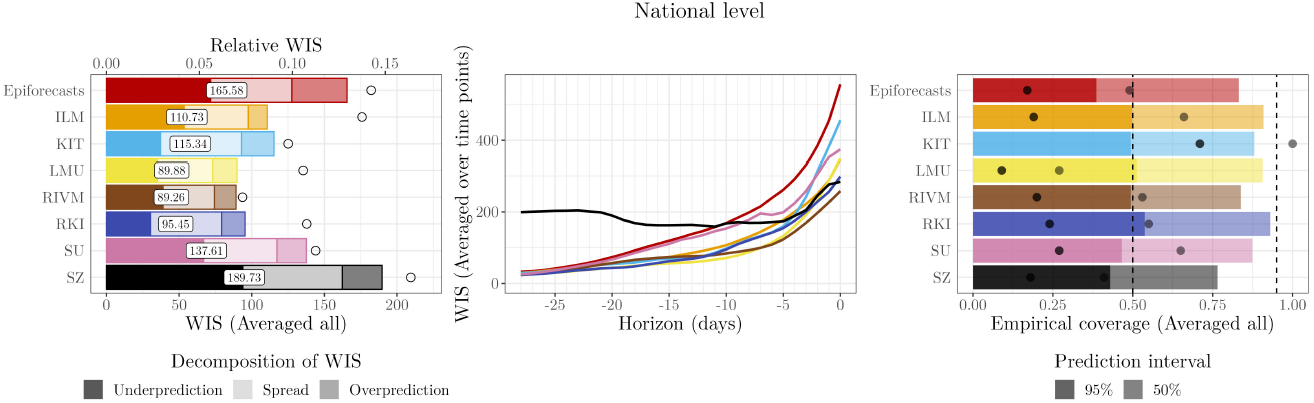
Performance of post-processed (`PP4`) individual-model nowcasts compared to the original versions, national level. Left: WIS (averaged over time points and horizons). Right: Coverage proportions (averaged over time points and horizons). In the left and right panel, circles (º) represent the results for the original models before post-processing, i.e., as in Figure 3.

We discuss results in more detail for the `LMU` and `SZ` models which, as mentioned in Section 4.2, have specific dispersion errors and biases. For `LMU`, we notice that the spread component of the WIS is larger than before, implying wider prediction intervals. We illustrate this for same-day nowcasts with *h* = 0 in Figure 5 (first row, left column; consider the respective panel of Figure 2 for comparison). The score improvements are consistent over nowcast horizons and dates (Figure 5, first row, middle and right columns). For the `SZ` model, although the overall WIS is not drastically improved, the underprediction component is much smaller and the coverage rates are better than before. As can be seen for nowcasts 14 days back in the second row of Figure 5, the post-processed `SZ` nowcasts no longer display a clear bias. The improvement in WIS values is pronounced for more distant horizons, while for short horizons there is actually a minor deterioration.

**Figure 5:**
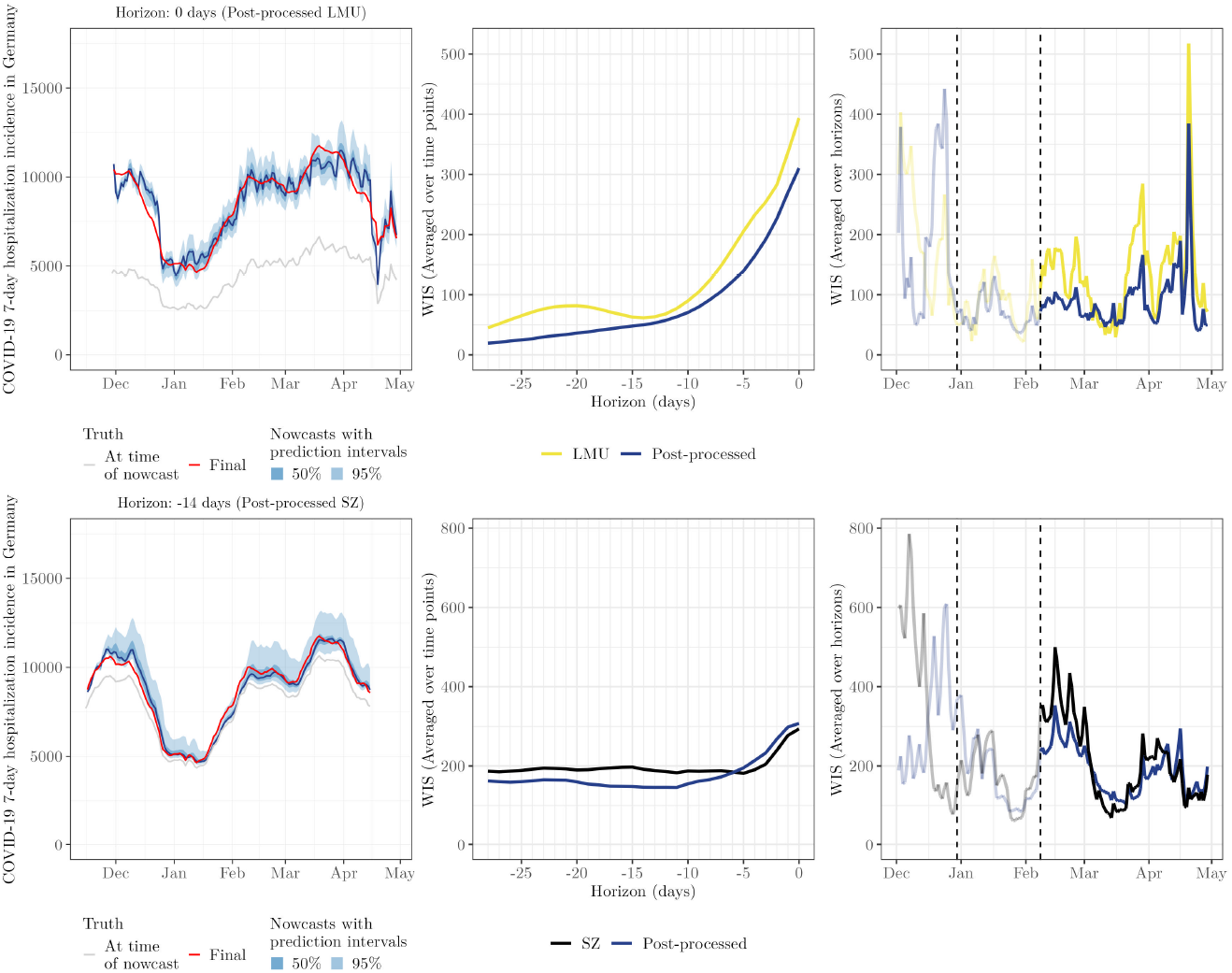
Left column: Same-day nowcasts for the post-processed `LMU` model (top) and nowcasts 14 days back for the post-processed `SZ` model (bottom). All nowcasts are at the national level and based on the post-processing scheme `PP4`. Middle column: Average WIS before and after post-processing, by nowcast horizon. Right column: WIS (averaged over horizons) before and after post-processing, per target date. The two dashed vertical lines represent December 30, 2021, i.e., the earliest target date, and February 8, 2022, i.e., the first nowcast date of the evaluation period. Scores before February 8 (greyed out) only partly enter into the reported average scores (with nowcasts referring to this period but issued on February 7 or before excluded).

For the other models (Supplementary Figures SF6–SF11), there are improvements in average WIS, but they are less consistent over time and nowcast horizons. This holds especially for the `KIT` model. As mentioned in Wolffram et al. (2023), the main shortcoming of the `KIT` model is an insufficient handling of weekday patterns, leading to different biases on different days of the week. This aspect cannot be corrected by our simple scaling approach.

### 4.4 Performance of ensemble approaches

We now turn to the performance of weighted nowcast ensembles. For the various approaches presented in Section 3.5, we again varied the way yet incomplete observations are used and whether parameters are shared across horizons; see the summary in the bottom part of Table 1. Note that due to extensive computing times, only a subset of approaches was applied to the stratified nowcasts (marked with a star symbol, ⋆). As before, we used a maximum value of *R* = 90 and assessed sensitivity to *R* = 60 and no upper limit on *R* (Figure SF13, Supplementary Material). The performance of the various combination approaches is summarized graphically in Figure 6 for the national level and Figure 7 for age strata and states. A graphical display of nowcasts produced by selected approaches is given in Figure 8. The results are discussed in subsections paralleling the structure of Section 3.5.

**Figure 6:**
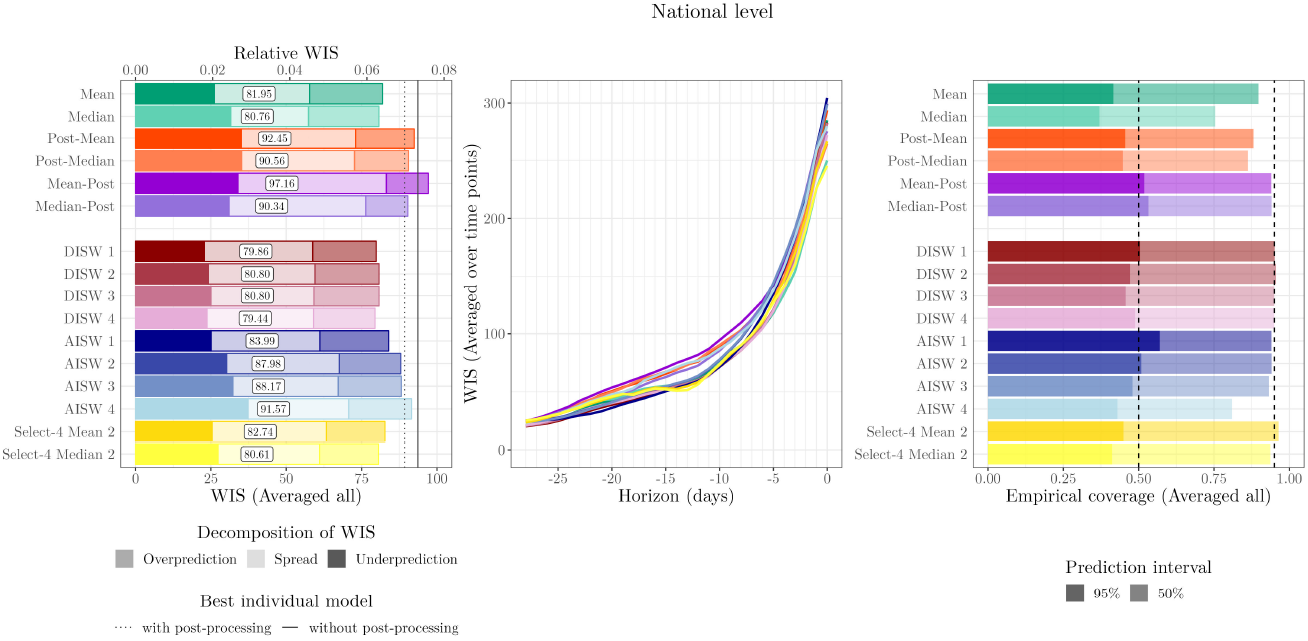
Performance of unweighted and weighted ensemble approaches at the national level. Left: WIS (averaged over time points and horizons). For reference, vertical lines indicate the performance of the best individual model with (dotted line) an without post-processing (solid line; in both cases RIVM). Middle: WIS (averaged over time points) by nowcast horizon. Right: Coverage proportions (averaged over time points and horizons).

**Figure 7:**
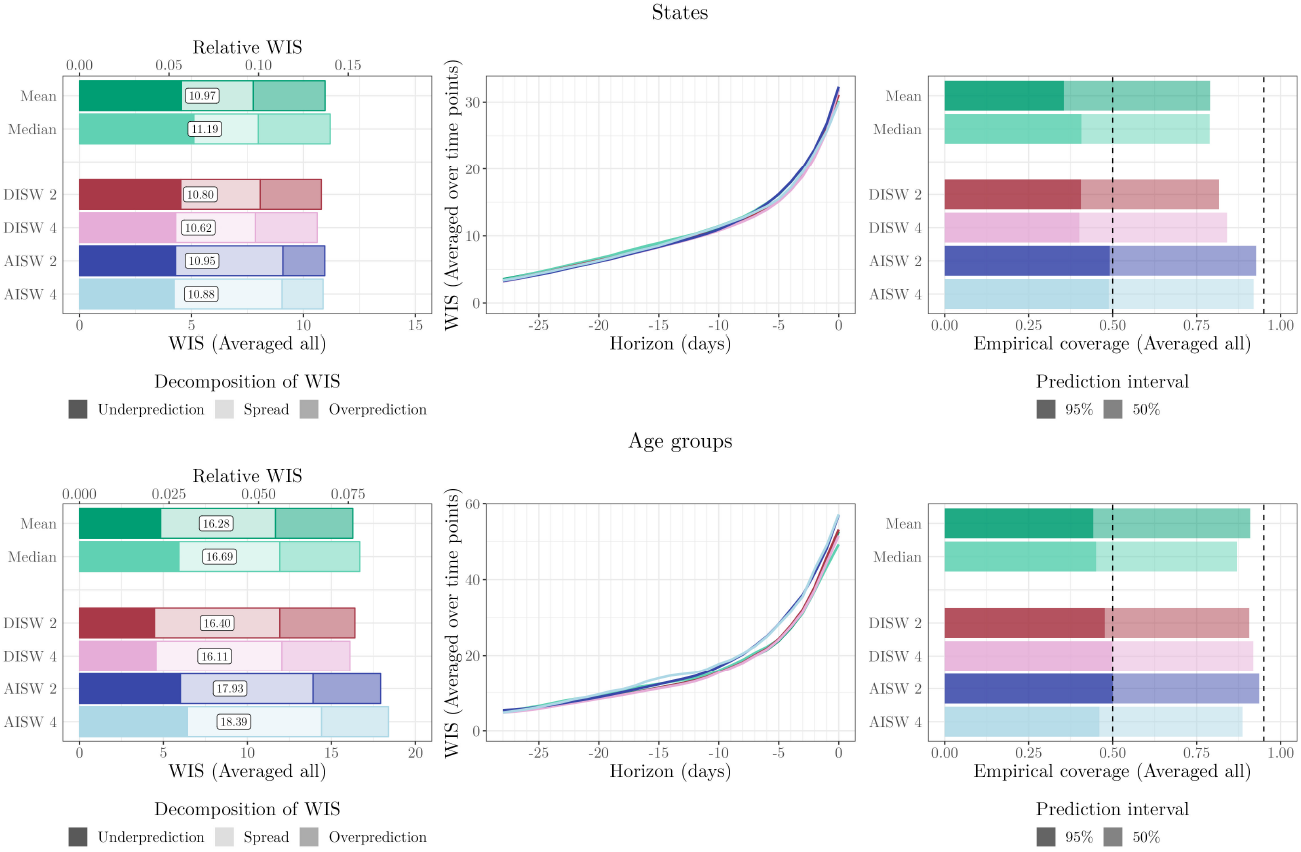
Performance of unweighted and weighted ensemble approaches at the state (top) and age-group levels (bottom; both averaged across strata). Left: WIS (averaged over time points and horizons). Middle: WIS (averaged over time points) by nowcast horizon. Right: Coverage proportions (averaged over time points and horizons). Note that due to extensive computing times, only a subset of approaches was applied to the stratified nowcasts (see Table 1).

**Figure 8:**
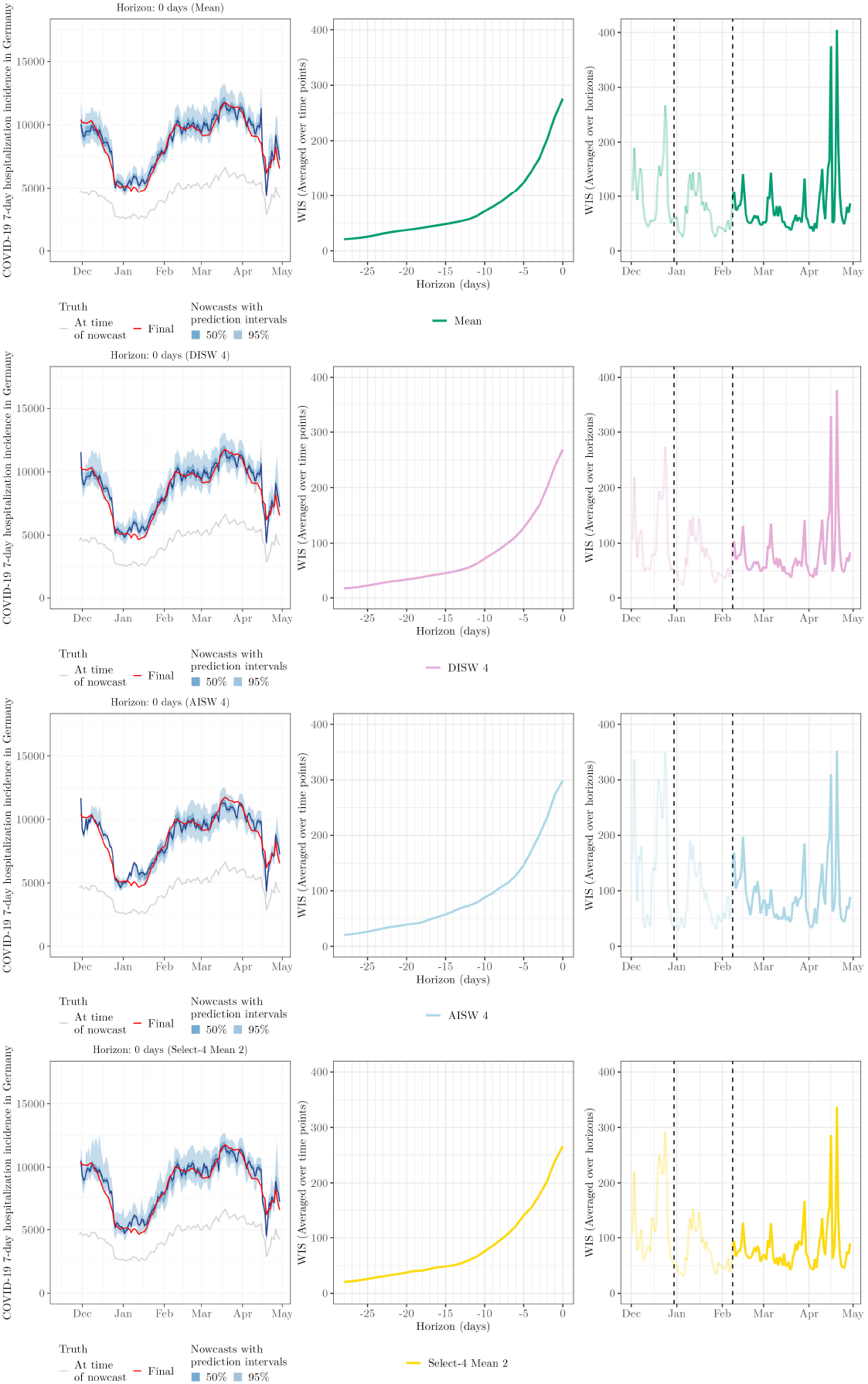
Illustration of same-day nowcasts for the `Mean`, `DISW4`, `AISW4` and `Select-4-Mean2` ensembles. See caption of Figure 5 for details on plot elements and Table 1 for details on the methods specifications.

#### 4.4.1 Unweighted ensembles

As already evoked in Section 4.2, the unweighted `mean` and `median` ensembles outperform all individual models in terms of average WIS, and most of them in terms of interval coverage. Even after post-processing (Section 4.3), the average WIS of all individual models remains inferior to the unweighted ensembles. For the following, the two unweighted ensembles can thus be seen as the baseline upon which more sophisticated combination approaches should improve.

#### 4.4.2 Post-processing-based approaches

The results achieved by unweighted averaging of post-processed nowcasts (`Post-Mean` and `Post-Median`) and post-processing of unweighted ensembles (`Mean-Post` and `Median-Post`) are similar, i.e., the order of post-processing and averaging does not seem to be decisive. In terms of interval coverage, both perform favourably. As can be seen from the WIS decomposition in the left panel of Figure 6, this is achieved by a widening of nowcast intervals (see the increased spread components). In terms of average WIS, however, the post-processing-based approaches are not only outperformed by the unweighted ensembles `mean` and `median`, but even some post-processed individual models. This is surprising given that post-processing improved the performance of all individual models.

While it is hard to provide any definitive explanation for the observed decrease in performance, one possible reason is that post-processing reduces the *diversity* of the ensemble. It is often argued that ensembles work best if their members are diverse and contribute distinct signals (DelSole et al., 2014).

By applying the same post-processing scheme to all members, or by glossing over the ensemble nowcast with a single post-processing method, characteristics of the post-processing method may dominate the ensemble characteristics, and diversity may be compromised. As illustrated in Figure SF12 (Supplementary Material), this is indeed the case in terms of pairwise approximate integrated quadratic distances between model nowcasts (see Supplementary Section SS3 on this metric). In the case of post-processing the unweighted ensembles, it is also possible that the margins for improvement by simple re-scaling are too modest in order to outweigh the cost of estimating scaling factors (see also Section 4.4.4).

#### 4.4.3 Direct inverse score weighting

The four considered variations of the direct inverse-score weighting overall perform similarly to the unweighted ensembles, with some modest improvements. The variant `DISW4` (weights varying over horizons, simple imputation) has the lowest average score, but by a margin that should not be interpreted as a meaningful difference. For the nowcasts stratified by age group and state, the results are overall similar, see Figure 7. As we will see in the following, the simple DISW approaches overall achieve the best performance of all considered combination approaches.

The uncertainty intervals of the DISW ensembles are somewhat wider than in the unweighted ensembles; consider again the spread components in the left panel of Figure 6 as well as the illustration of nowcasts in Figure 8. This results in improved calibration at the national and age group levels. Apart from this, however, the DISW forecasts look quite similar to the unweighted `mean` nowcasts.

The weights assigned to the different models are quite close to uniform for the predictive median, see the middle panel of Figure 9. For the 0.025 and 0.975 quantiles, weights are more imbalanced and vary over time. The `RIVM` model, which tends to over-predict (see WIS decomposition in Figure 3), receives little weight for the 0.025 quantile. The `LMU` model, on the other hand, receives little weight for the 0.975 quantile, as it tends to underpredict. This explains the aforementioned widening of prediction intervals.

**Figure 9:**
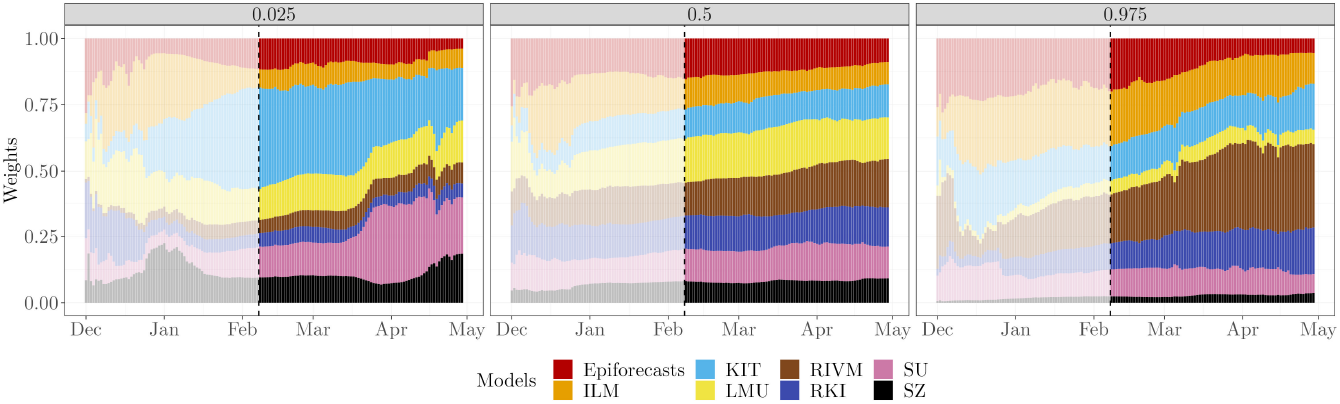
Estimated weights for the 0.025, 0.5, and 0.975 quantiles based on the direct inverse-score weighting method `DISW2` (weights shared across horizons, simple imputation). Weights are shown for the national level. As in Figure 5, results preceding the actual evaluation period are greyed out.

To illustrate the behaviour when weights are only based on few historical nowcasts and observations, we also display the initial period November 29, 2021, through February 7, 2023 (greyed out), which is excluded from the evaluation. As could be expected, the weights fluctuate more strongly during this period.

#### 4.4.4 Adjustable inverse score weighting

We now turn to the AISW method, which unlike the DISW approach requires determining scaling and weighting parameters based on past pairs of nowcasts and observations. In practice, this resulted in considerably increased computational effort, but did not translate to gains in performance in terms of average WIS. While the difference to the unweighted and DISW ensembles is not drastic, it is consistent across specifications 1 through 4. The interval coverage rates are similar to those of DISW.

Figure 10 shows the estimated weights for setting `AISW2`. The corresponding plots for the other AISW settings, along with the estimated weights aggregated by horizon or quantiles (where applicable), are presented in the Supplementary Material. Several observations can be made from Figure 10. Firstly, the weights are less smooth over time than in Figure 9. In some instances, e.g., in early March for the 0.025 quantile, there are small jumps, which may indicate the presence of several local optima in the objective function (note that our grid search ensures that we do not end up in a local optimum, but the global optimum can “jump” to a different local optimum from one day to the other). For the 0.025 quantiles, the effective model weights (i.e.,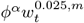) sum up to a value below one. The scaling parameter *ϕ*^*α*^ is thus below one and leads to lower (more conservative) ensemble quantiles. For the predictive median, almost no re-scaling takes place, while for the 0.975 quantile there is likewise some downscaling. Compared to Figure 9, the differences between weights received by different models are exacerbated, i.e., the AISW ensemble emphasizes models with better historical WIS values even more (meaning that the *θ*^*h,w*^ exceed one). This is especially pronounced for the 0.975 quantile, where the RIVM model receives a large weight towards the end of our study period.

**Figure 10:**
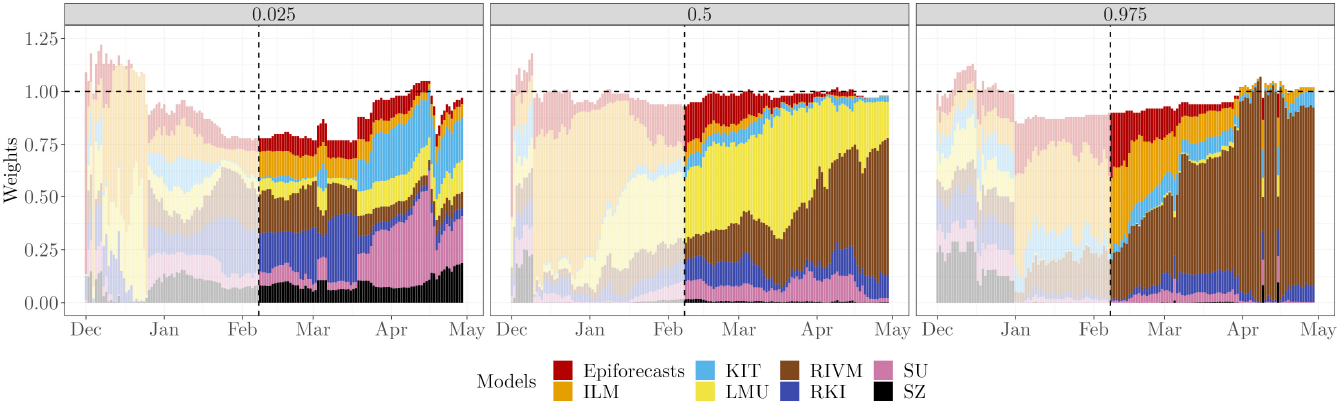
National-level weights for the 0.025, 0.5, and 0.975 quantiles based on the `AISW 2` method (weights and scaling parameter shared across horizons, simple imputation). Due to the introduced scaling parameter *ϕ*^*α*^, the weights are not required to sum up to 1. The horizontal dashed line represents `weight` = 1.

For nowcasts stratified by states and age groups (Figure 7), the performance of the AISW approach is somewhat more favourable. For state-level nowcasts, in which case 16 times more data are available to determine the weights in a data-driven way, the AISW achieves minimally better scores than the unweighted ensemble and minimally worse than the DISW. For age groups, in which case 6 times more data are available, the AISW ensembles again fall behind the unweighted and DISW variations.

The results at the national and stratified levels indicate that the estimation of weighting parameters may come at the cost of fluctuating and somewhat unstable ensemble weights. The fluctuating nature of the weights may either indicate that there is not enough data to estimate them reliably, or that there is not actually a temporally stable “right” configuration of weights.

#### 4.4.5 Top-*n* model selection

Lastly, we consider the ensembles based on selection rather than weighting of members. As the user needs to specify the number *n* of maintained models in advance, we assess the performance for all values *n* = 1, …, 8 (with *n* = 1 corresponding to the selection of the top model only, and *n* = 8 corresponding to the unweighted ensemble).

In Figure 6, we show the results for *n* = 4, i.e., at each time point the better half of the models (over the training period) is included in the ensemble, with selection performed separately per horizon (`Select-4-Mean2` and `Select-4-Median2`). A graphical illustration of the respective nowcasts has been included in the bottom row of Figure 8. Despite some visually discernible differences to the unweighted ensembles (top panel), the average WIS values of `Select-4-Mean2` remain very close to those of the unweighted ensemble. Interval coverage rates are again somewhat improved. Figure 11 shows the overall WIS for the different values of *n* = 1, …, 8 and the mean (left panel) and median (right panel) as the combination function. Red dots represent the results when the set of *n* models is updated every day, as would be done in a real-time application. For context, we show the results for all possible combinations of *n* models, keeping the selection of models constant over time, horizons and quantiles. Several conclusions can be drawn from the plot. Firstly, performance overall improves the more models are included into the ensemble, and only few model combinations at *n* = 3 through 7 achieve slight improvements over the full ensemble with *n* = 8. On the other hand, selection in real time (red dots) is always quite close to the optimum that could be achieved with a time-constant model selection, and comes close to the full unweighted ensemble from *n* = 3 onwards.

**Figure 11:**
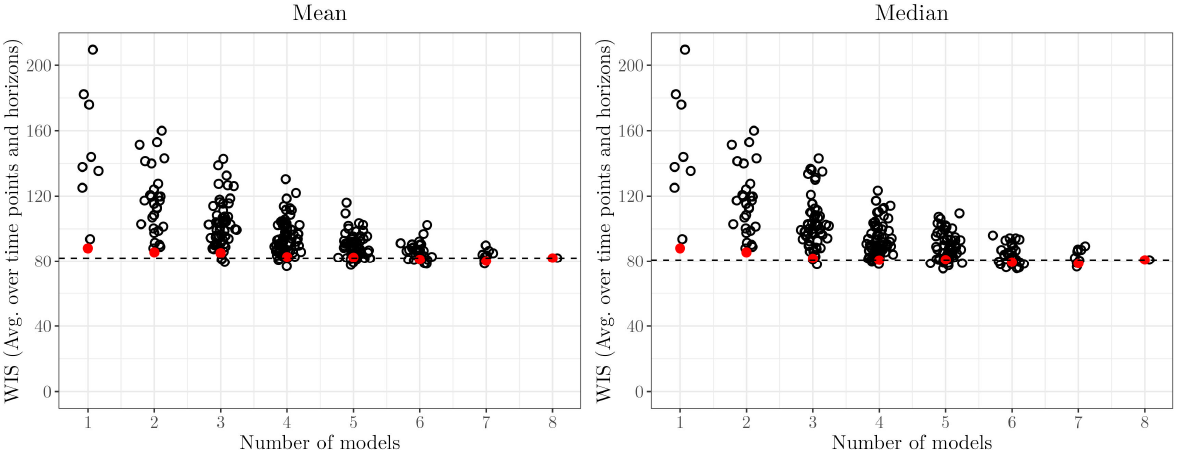
WIS (averaged over time points and horizons) for *n* = 1, …, 8 in the `Select-n-Mean2` (left) and `Select-n-Median2` (right) models. Red circles show results for model selection updated each day, as would be done in a real-time setting. For context, black circles show average values for all possible combinations of models when keeping the selection fixed over time. The horizontal dashed line represents the average WIS achieved by the full ensemble with all eight member models.

In the Supplementary Material, we present the corresponding results for the settings where the models are chosen jointly for all horizons (`Select-n-Mean1` and `Select-n-Median1`). Performance is overall somewhat weaker than when selection is done separately per horizon.

While again there is no clear improvement over the unweighted ensemble, our results indicate that the effort necessary to maintain an ensemble model with numerous members may be reduced by restricting it to a few well-chosen members after an initial performance assessment.

## 5 Discussion

In this paper, we proposed and analyzed different post-processing and ensemble techniques for the nowcasting of infectious diseases. In an application to COVID-19 hospitalization numbers from Germany, we found that post-processing of individual models yielded performance gains across almost all considered models and technical specifications. This held both in terms of average WIS values and nowcast interval coverage. In this setting, we also found benefits in our proposed approaches to include yet incomplete data points into the fitting of the post-processing model. Somewhat surprisingly, post-processing of unweighted ensemble nowcasts did not yield improved performance, nor did post-processing of members prior to ensembling. More generally, it proved very challenging to improve upon unweighted mean and median ensembles. A straightforward direct inverse-score weighting approach led to very minor improvements, while a more sophisticated approach with weights optimized based on recent nowcast and observation pairs actually led to a decline in performance. Data-driven restriction of the ensemble to models with good recent performance did not yield improved performance either. On a more positive note, the results indicate that the size of the ensemble, and thus the effort needed to maintain it, can be reduced without major losses in performance.

In the present paper we attempted to cover a spectrum of methods of moderate complexity which could be employed in practice. Many other extensions and alternative variations could be explored (e.g., we did not attempt to weight post-processed member models). However, our general takeaway is that added complexity did not translate to improved performance. Some more flexible approaches we explored, such as quantile regression with unconstrained weights for each model, proved to be intractable in our setting. To overcome this, most of our approaches assumed that successful individual models should receive increased weight, but this may not necessarily be the case. Other approaches to addressing collinearity issues, such as clustering models into a small number of sufficiently distinct groups, could be explored. Another promising avenue involves using machine learning methods that can capture complex dependency structures while effectively counteracting overfitting.

More or less sophisticated weighting schemes being unable to outperform simple unweighted ensembles is a common finding in the literature, and Stock and Watson (2004) have coined the term “forecast combination puzzle” for this phenomenon. Various theoretical and empirical arguments have been brought forward to explain it (e.g., Claeskens et al. 2016; Smith and Wallis 2009). The essence of these is that estimated weights are often poorly identified and quite variable. This has a negative effect on performance, which may exceed the cost of the bias inherent in uniform weighting. Estimation of weights is thus less promising the closer the “true” weights are to uniformity.

A number of limitations of the present study need to be acknowledged. All our analyses were conducted retrospectively rather than in real time. This introduces the risk of hindsight bias and enabled us to explore approaches of higher computational cost than might have been feasible in real time. Also, the evaluation period spans only roughly 12 weeks, and early on the number of forecast and observation pairs available for training purposes was rather low. It is possible that trained ensembles would work better with more training data available (though it is not clear to which degree “old” training data will help improve nowcasts).

We moreover simplified our task in some respects and ignored a few challenges which may arise in a real-time application. Firstly, occasional faulty submissions of individual models would need to be caught in operational use as they can strongly perturb weighted mean ensembles (we note that median ensembles are more robust, but lend themselves less to weighting). Similarly, missing submissions are not addressed. The considered post-processing and combination methods were chosen such that they can relatively easily be extended to account for missing submissions (see Ray et al. 2023), but it is unclear how this will affect the performance of the ensemble.

Concerning the post-processing scheme, we note that our methods are unable to correct some shortcomings of the original nowcasts which are easy to spot for the human observer. Notably, the issues of the `KIT` model related to weekday effects went uncorrected in our scaling approach. Consequently, it was of little use to improve the `KIT` nowcasts.

The fact that improved calibration (interval coverage) of post-processed and weighted ensembles did not yield improved performance in terms of average WIS may also reflect that this score is relatively insensitive to overconfident predictions (see discussion in Bracher et al. 2021a). It would have been desirable to apply also other scores like the logarithmic score which is known to penalize dispersion errors more severely. However, this was not feasible due to the quantile-based format in which nowcasts were collected.

## Declarations

### Conflict of interest

The authors declare that they have no conflict of interest.

### Data availability statement

The nowcast data for all individual models are available at https://github.com/KITmetricslab/hospitalization-nowcast-hub. The code used to reproduce the results presented throughout this paper is available at https://github.com/avramaral/ensemble_learning.

## Acknowledgements

The authors would like to thank all contributors to the German COVID-19 Hospitalization Now-cast Hub. André Victor Ribeiro Amaral acknowledges support from Karlsruhe Institute of Technology via the Aspirant Postdoc Grant. Daniel Wolffram and Johannes Bracher were supported by the German Federal Ministry of Education and Research (BMBF) via the project RESPINOW. Daniel Wolffram was moreover supported by the Helmholtz Association under the joint research school HIDSS4Health— Helmholtz Information and Data Science School for Health. Johannes Bracher was moreover supported by the German Research Foundation (DFG), project 512483310.

## Notes

### Competing Interest Statement

The authors have declared no competing interest.

### Author Declarations

The nowcast data for all individual models, as well as the true data, are available at https://github.com/KITmetricslab/hospitalization-nowcast-hub

### Summary of Updates

In this updated version, we revised the entire manuscript (correcting typos, rephrasing slightly confusing sentences, adding missing details, etc.) and conducted additional simple analyses (e.g., age stratification of results for individual models and diversity of nowcasts before and after post-processing--all included in the Supp. Material). Additionally, the Appendix was moved to the Supp. Material.

