## Supplementary Material for "Post-processing and weighted combination of infectious disease nowcasts"

### SS1 Individual Models

A summary of characteristics of the eight individual models can be found in (Wolffram et al., 2023, Table 1). We here only provide a high-level summary for each model, along with the reference to the respective original paper or documentation.

1. `Epiforecasts` (Abbott et al., 2021): A Bayesian approach combining flexible modelling of delay distributions (including weekday effects) and a random walk prior on the latent time series of complete hospitalization counts.

2. `ILM` (Heyder and Hotz, 2021): Predictions of yet unreported hospitalizations are based on case counts, for which suitable multiplication factors are derived per age group. Uncertainty intervals are based on past nowcast errors. Note that we here do not use the model as run in real time, but a retrospectively re-run version with an assumed maximum delay of 42 days. The real-time model featured a maximum delay of 80 days, which was not suitable for the purposes of the present study.

3. `KIT` (Wolffram et al., 2023, Supplementary Section E): A simple multiplication factor model ignoring weekday effects. Uncertainty intervals are based on past nowcast errors.

4. `LMU` (Schneble et al., 2021): A frequentist nowcasting approach combining a generalized additive model for the reporting triangle and a sequential multinomial model for the delay distribution.

5. `RIVM` (van de Kassteele et al., 2019): The reporting triangle is modelled using a bivariate spline surface, accounting for weekday effects. This is used to extrapolate to the unobserved parts of the triangle.

6. `RKI` (an der Heiden and Hamouda, 2020): Logistic regression is used to model conditional reporting probabilities, taking into account various covariates (weekdays, states, age groups, etc.).

7. `SU` (Günther et al., 2021): Conceptually close to Epiforecasts, combines a random walk prior with a discrete-time hazard model for the reporting delay.

8. `SZ`: Nowcasts are based on empirical ratios of preliminary and completed data, for which various quantiles are computed.

#### SS2 Decomposition of the weighted interval score

Rather than via the linear quantile score, the weighted interval score can also be defined as a weighted sum of interval scores (thus its name). The interval score for a prediction interval $[l_\beta, u_\beta]$ at nominal coverage level $(1-\beta)$ is given by (Gneiting and Raftery, 2007)

$$\text{IS}_\beta(l_\beta, u_\beta, x) = (u_\beta - l_\beta) + \frac{2}{\beta} \times (l_\beta - x) \times \mathbb{1}(x < l_\beta) + \frac{2}{\beta} \times (x - u_\beta) \times \mathbb{1}(x > u_\beta).$$

It obviously consists of three components, namely

1. a component for forecast dispersion, given by the interval width $u_\beta - l_\beta$.
2. a component for overprediction, i.e. a penalty in the case $x > u$.
3. a component for underprediction, i.e., a penalty in the case $x < l$.

Now assume that $A$ is uneven and that the quantile levels are chosen such that $q^{\alpha_1}, \dots, q^{\alpha_A}$ correspond to the predictive median $m = q^{\alpha_{(A+1)/2}}$ and the ends $l_{\beta_1} = q^{\alpha_1}, \dots, l_{\beta_{(A-1)/2}} = q^{\alpha_{(A-1)/2}}, u_{\beta_1} = q^{\alpha_{(A+1)/2+1}}, \dots, u_{\beta_{(A-1)/2}} = q^{\alpha_A}$ of $(A-1)/2$ nested central prediction intervals. These intervals have nominal coverage levels $(1-\beta_j) = \alpha_{A+1-j} - \alpha_j, j = 1, \dots A$. The weighted interval score can then also be written as (Bracher et al., 2021)

$$\text{WIS}(l_{\beta_1}, \dots, l_{\beta_{(A+1)/2}}, m, u_{\beta_{(A+1)/2}}, \dots, u_{\beta_1}, x) = \frac{1}{A} \times \left( |x - m| + \sum_{j=1}^{(A-1)/2} \beta_j \times \text{WIS}(l_{\beta_j}, u_{\beta_j}, x) \right). \quad (1)$$

It thus inherits the decomposition of the interval score. In practice, the components can also be computed in a more straightforward way without recurring to the somewhat involved Equation (1). This can be done as follows

1. The spread component equals the WIS with the observation $x$ replaced by the predictive median $m$.
2. The overprediction component is zero if $x < m$. If $x > m$, it is given by the difference of the WIS and the spread component.
3. Accordingly, the underprediction component is zero if $x > m$. If $x < m$ it is given by the difference of the WIS and the spread component.

#### SS3 The approximate integrated quadratic distance

The simple imputation approach, where preliminary observations are replaced by up-to-date predictive medians in forecast evaluation, neglects that there is uncertainty attached to these imputed values. It would be desirable to account for this and take into account the full current predictive distribution (or, in our setting, all available quantiles). To this end we use an approximation of the integrated quadratic distance (Thorarinsdottir et al., 2013), which we motivate below.

As mentioned in the main manuscript, the WIS is a quantile-based approximation of the continuous ranked probability score (CRPS). For a predictive cumulative distribution function $F$ and observed value $x$ this score is defined as

$$\text{CRPS}(F, x) = \int_{-\infty}^{\infty} [F(z) - \mathbb{1}(z \geq x)]^2 dz.$$

For the CRPS, a setting similar to ours is mentioned by Friederichs and Thorarinsdottir (2012). They address the evaluation of forecasts for quantities observed with an observation error. Denoting by $x$ the observed value including the observation error, they propose to use the score (see also Brehmer and Gneiting (2019))

$$S(F, x) = \int_{-\infty}^{\infty} [F(z) - \Phi(z - x)]^2 dz,$$

where $\Phi$ is the cumulative density function assumed for the observation error. This can be read as a comparison of two probability density functions: the one of the forecast, and an assumed distribution of the true value given the imperfect observation $x$. The resulting distance between the two cumulative distribution functions is called the *integrated quadratic distance* (IQD) (Thorarinsdottir et al., 2013).

This fits our setting well, as we likewise wish to compare one predictive density to an "uncertain" observation described by another probability distribution. Denoting the CDF of the nowcast distribution to evaluate by $F$ and the CDF of the most recent nowcast used as the "uncertain truth" by $F^*$, we could therefore use the integrated squared distance

$$\text{IQD}(F, F^*) = \int_{-\infty}^{\infty} [F(z) - F^*(z)]^2 dz$$

for evaluation. This, of course, is not feasible in practice as we only know a set of predictive quantiles for both $F$ and $F^*$. We denote these quantiles by $q_F^{\alpha_1}, \dots, q_F^{\alpha_A}$ and $q_{F^*}^{\alpha_1}, \dots, q_{F^*}^{\alpha_A}$, respectively. It can be demonstrated that an approximation of the IQD paralleling the approximation of the CRPS by the WIS is (Resin et al., 2024)

$$\text{IQD}(F, F^*) \approx \sum_{i=1}^{A} \sum_{j=1}^{A} \frac{\alpha_{i+1} + \alpha_{i-1}}{2} \times \frac{\alpha_{j+1} + \alpha_{j-1}}{2} \times \chi(\alpha_i, \alpha_j, q_F^{\alpha_i}, q_{F^*}^{\alpha_j}) \times \left| q_F^{\alpha_i} - q_{F^*}^{\alpha_j} \right|,$$

where

$$\chi(\alpha_i, \alpha_j, q_F^{\alpha_i}, q_{F^*}^{\alpha_j}) = \begin{cases} 1 & \text{if } (\alpha_i > \alpha_j \text{ and } q_F^{\alpha_i} < q_{F^*}^{\alpha_j}) \text{ or } (\alpha_i < \alpha_j \text{ and } q_F^{\alpha_i} > q_{F^*}^{\alpha_j}) \\ \frac{1}{2} & \text{if } \alpha_i = \alpha_j \\ 0 & \text{else.} \end{cases}$$

We thus use this expression to evaluate nowcast quantiles against preliminary observations accounting for the remaining uncertainty. When evaluating the performance separately per quantile level $\alpha_i$ of $F$, we can just use the summands for that $\alpha_i$, just like we are only using the respective linear quantile score rather than the full WIS if the observed value $x$ is known with certainty.

We note that an intuitive way of thinking of this procedure is the following: The approximate IQD is essentially (up to a typically small additive constant) equivalent to the expected $\text{WIS}(F, y)$ if we assume that $y$ comes from the latest nowcast distribution $F^*$. If nowcasts were available as full probability distributions, there would be an exact mathematical equivalence. In our practical setting, the fact that only quantiles of $F^*$ are available makes the described approximations necessary, but the general principle remains the same.

#### SS4 Age stratification of results for individual models

In this section, we present a complementary figure, along with comments for interpretation, showing the (relative) WIS stratified per age group for all individual models and the unweighted ensemble. See also Section 4.2.

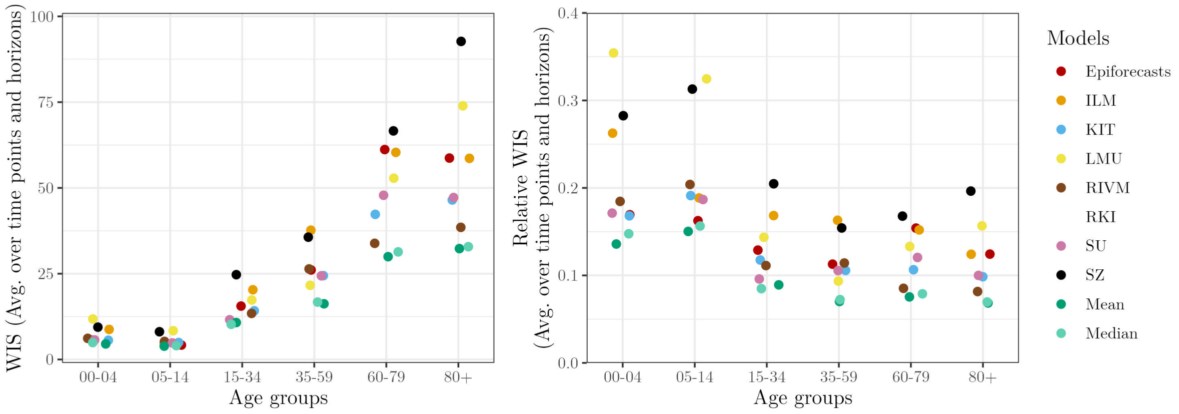


Figure SF1: Model performance of original models and unweighted ensembles from Wolffram et al. (2023) stratified by age groups. Left: WIS (averaged over time points and horizons). Right: Relative WIS (averaged over time points and horizons) with respect to a naïve baseline of no delay correction.

As illustrated in Figure SF1 (left panel), average WIS values are larger for the older age groups. This, however, merely reflects the scale-dependence of the WIS mentioned in Section 4.2, and the fact that most hospitalizations occur among older persons. Once moving to the relative WIS scale (right panel), the picture flips and the strongest improvements over the baseline are achieved in the older age groups. This may reflect the fact that higher-count outcomes are usually less noisy, and slightly easier to predict on a relative scale; see (Bosse et al., 2023, Figure 7) for a similar empirical result.

Another important takeaway is that the performance ranking of different models remains fairly constant across age groups (with the ensembles achieving the lowest scores and SZ typically receiving slightly higher scores than the other individual models).

### SS5 Results of post-processing nowcasts

In this Section, we present the supplementary results based on the post-processing methods implemented in Section 4.3.

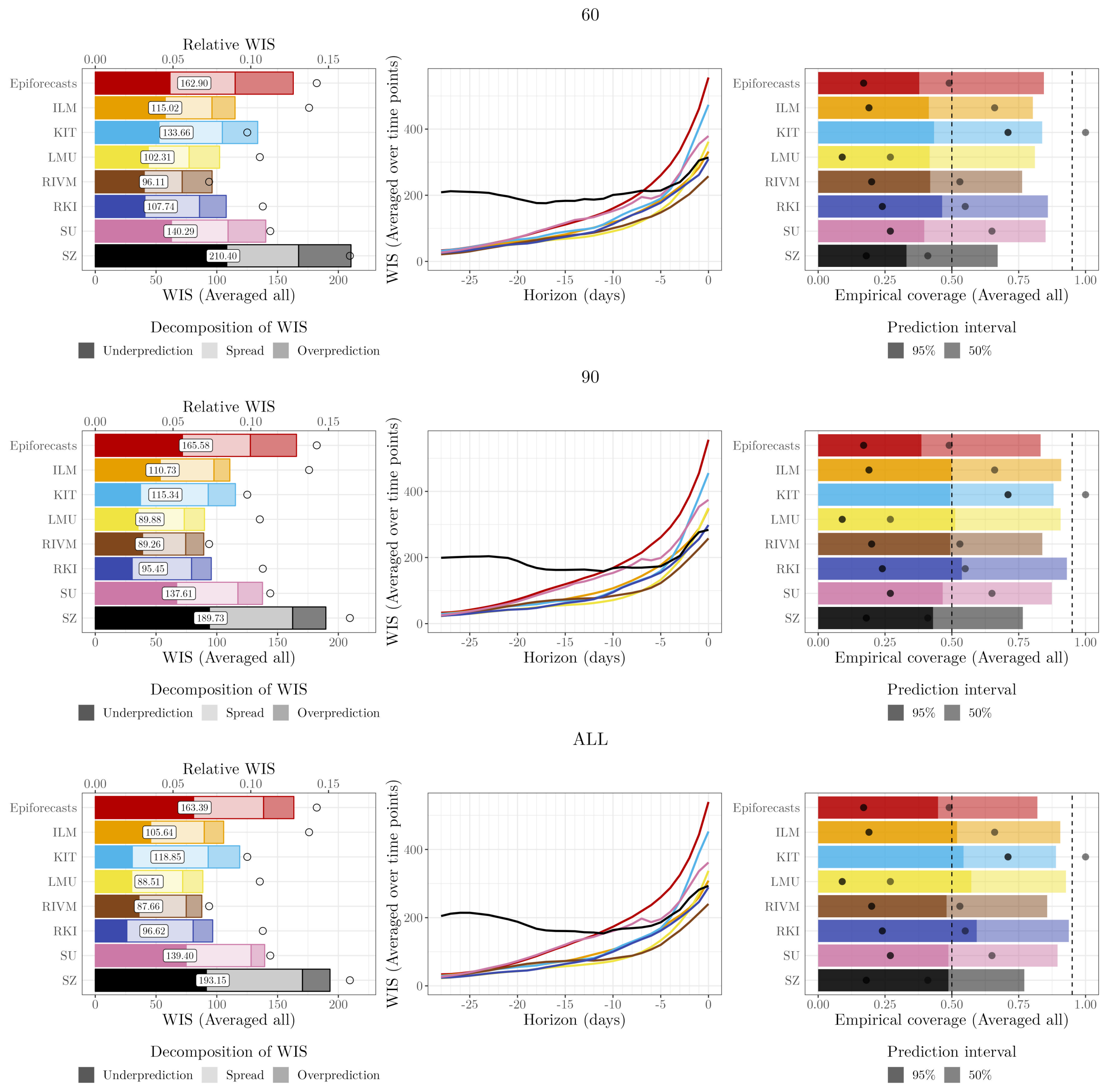


Figure SF2: Sensitivity of post-processing results from Figure 6 to the choice of maximum value $R$ of historical nowcast dates used in the score minimization. The middle row shows $R = 90$ as in Figure 4 of the main manuscript. The top and bottom rows show results for $R = 60$ and no upper limit $R$, respectively. See caption of Figure 4 for details on plot elements.

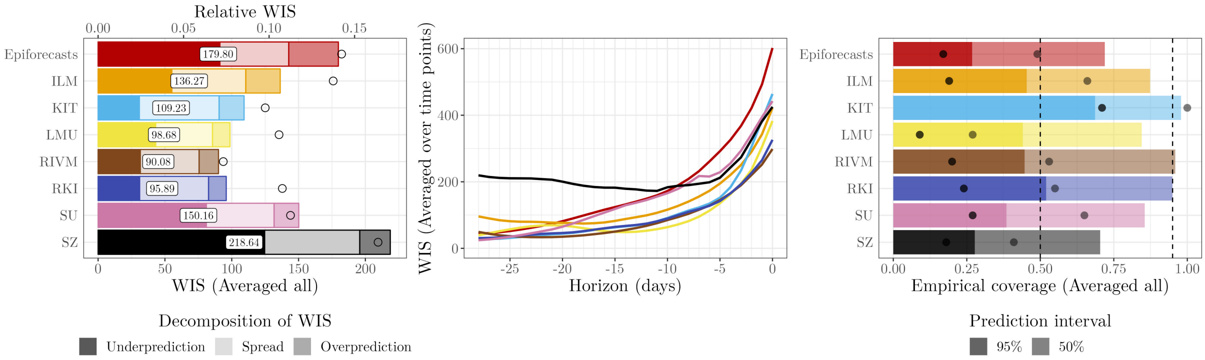


Figure SF3: Postprocessing results for setting PP1, i.e., scaling parameter shared across horizons while discarding incomplete observations. See caption of Figure 4 for details on plot elements.

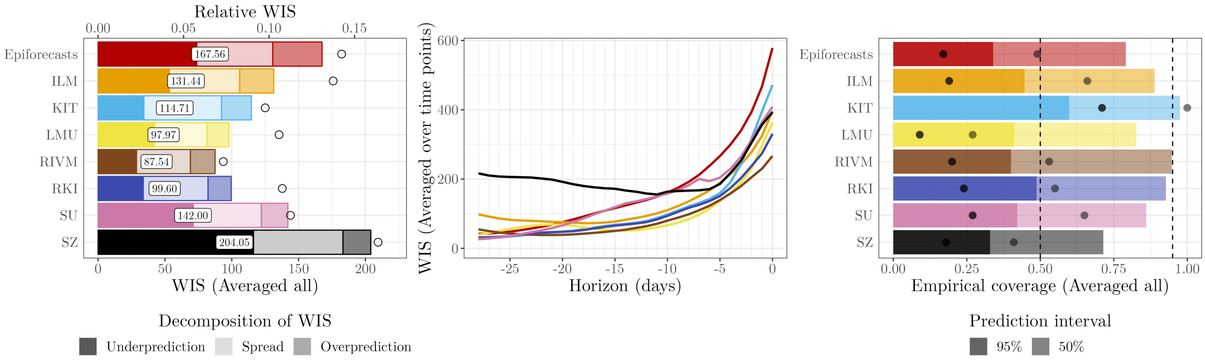


Figure SF4: Postprocessing results for setting PP2, i.e., scaling parameter shared across horizons, simple imputation. See caption of Figure 4 for details on plot elements.

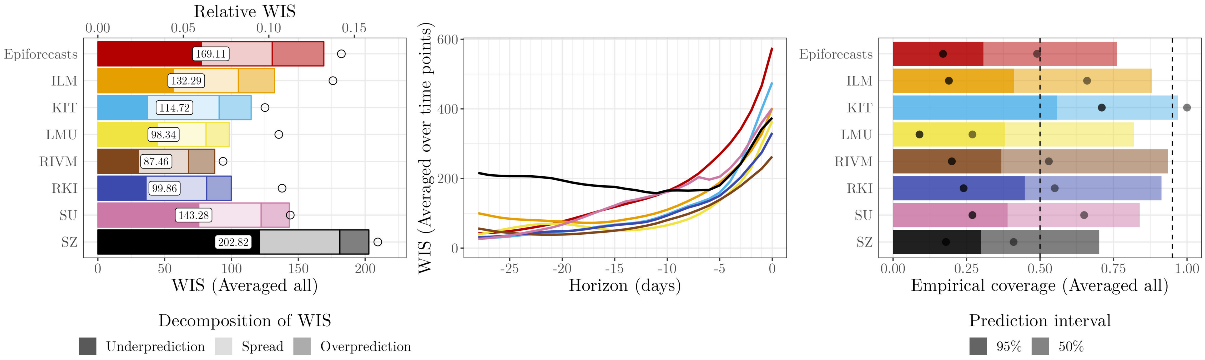


Figure SF5: Postprocessing results for setting PP3, i.e., scaling parameter shared across horizons, imputation with uncertainty. See caption of Figure 4 for details on plot elements.

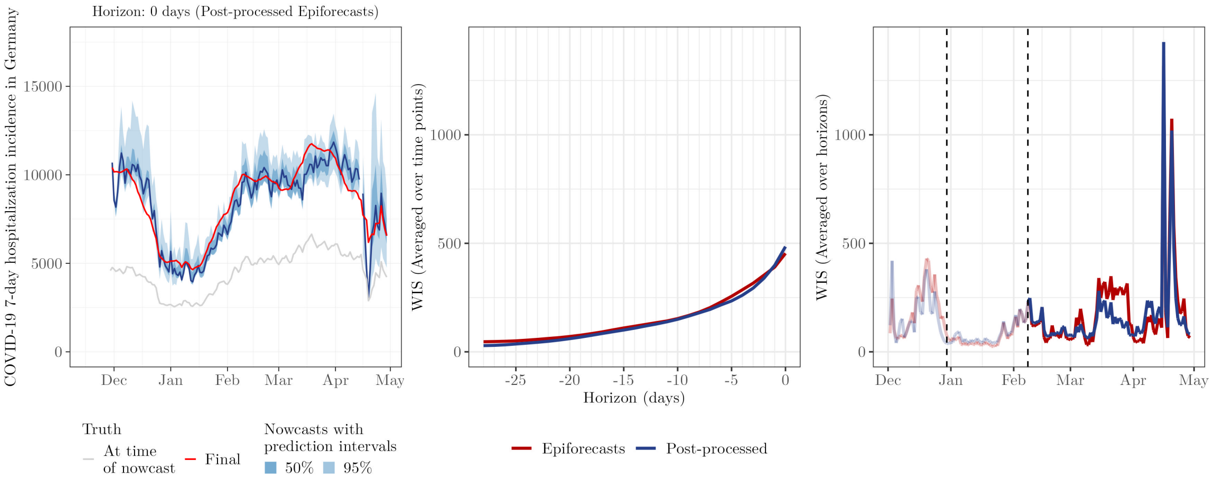


Figure SF6: Graphical display and performance of same-day nowcasts from the post-processed `Epiforecasts` model (`PP4`). See Figure 5 for details on the plot elements.

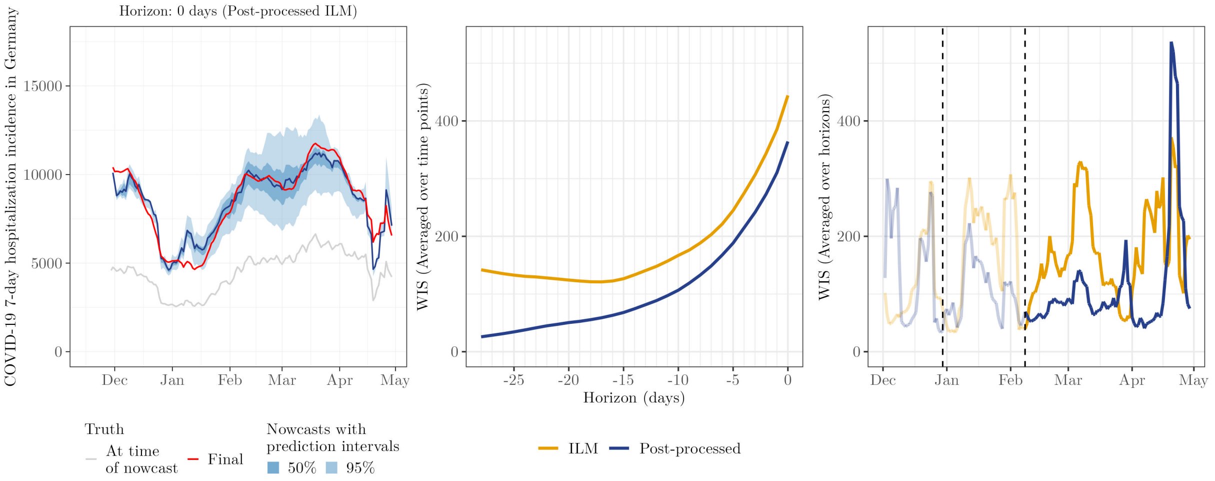


Figure SF7: Graphical display and performance of same-day nowcasts from the post-processed `ILM` model (`PP4`). See Figure 5 for details on the plot elements.

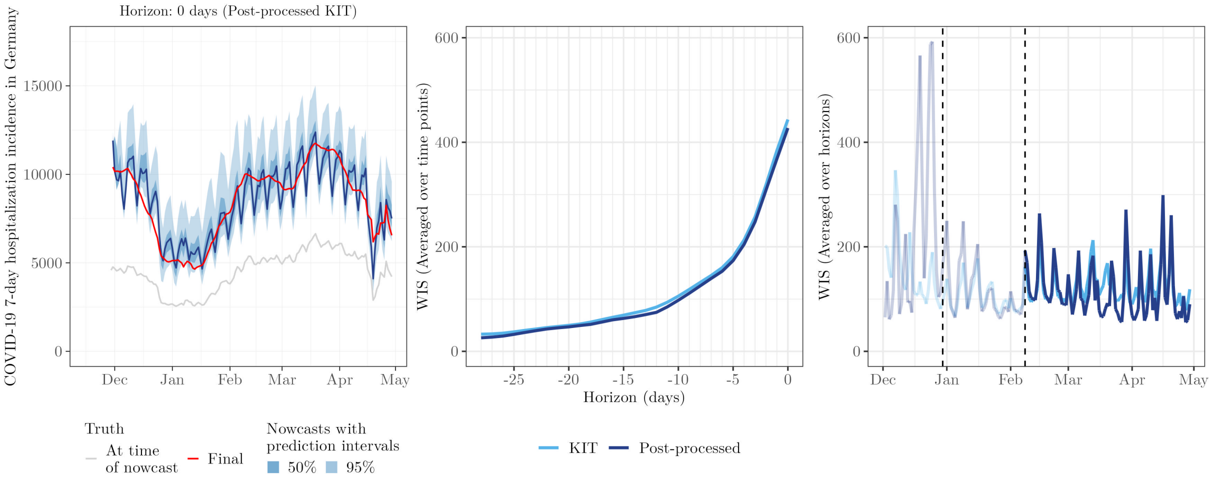


Figure SF8: Graphical display and performance of same-day nowcasts from the post-processed `KIT` model (`PP4`). See Figure 5 for details on the plot elements.

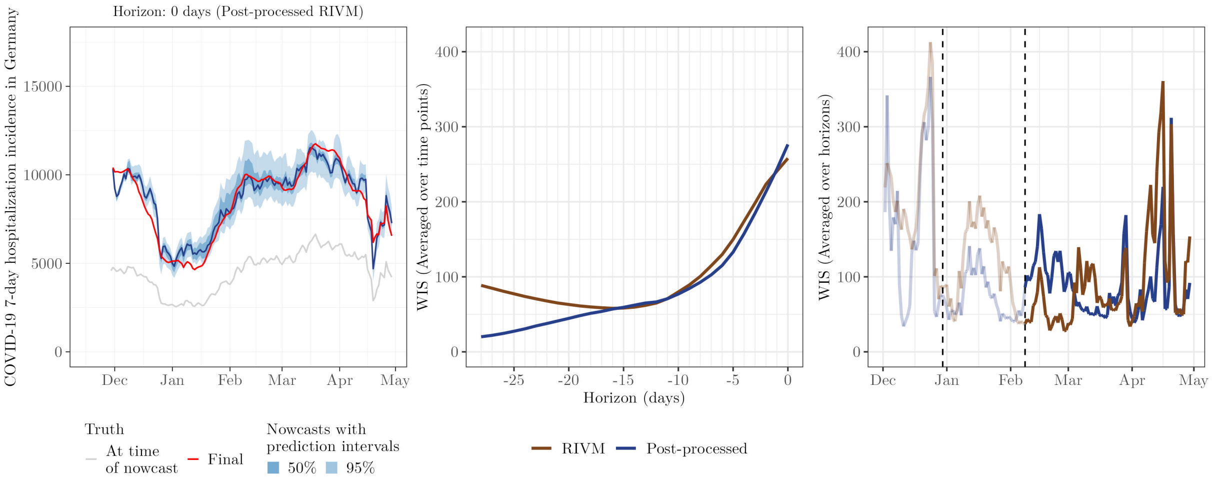


Figure SF9: Graphical display and performance of same-day nowcasts from the post-processed `RIVM` model (`PP4`). See Figure 5 for details on the plot elements.

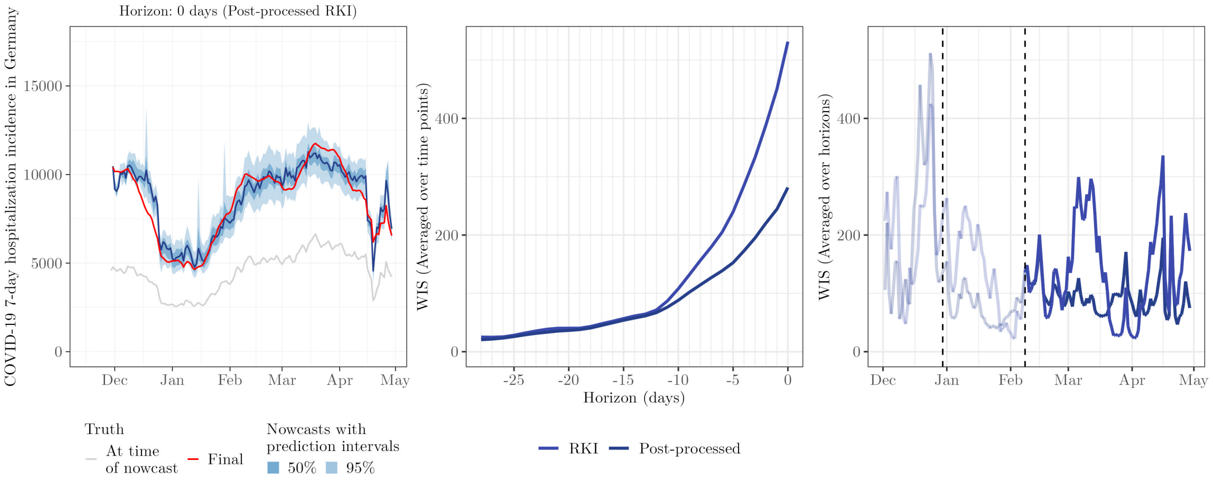


Figure SF10: Graphical display and performance of same-day nowcasts from the post-processed `RKI` model (`PP4`). See Figure 5 for details on the plot elements.

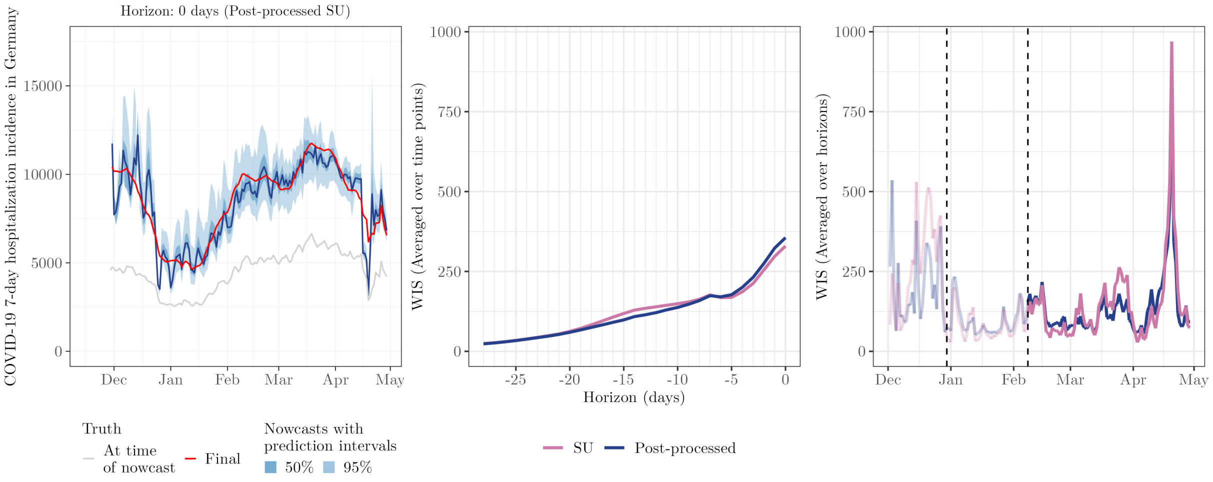


Figure SF11: Graphical display and performance of same-day nowcasts from the post-processed `SU` model (`PP4`). See Figure 5 for details on the plot elements.

#### SS6 Diversity of nowcasts before and after post-processing

In this section, we present an additional figure regarding the ensemble of post-processed nowcasts, as described in Section 4.4.2.

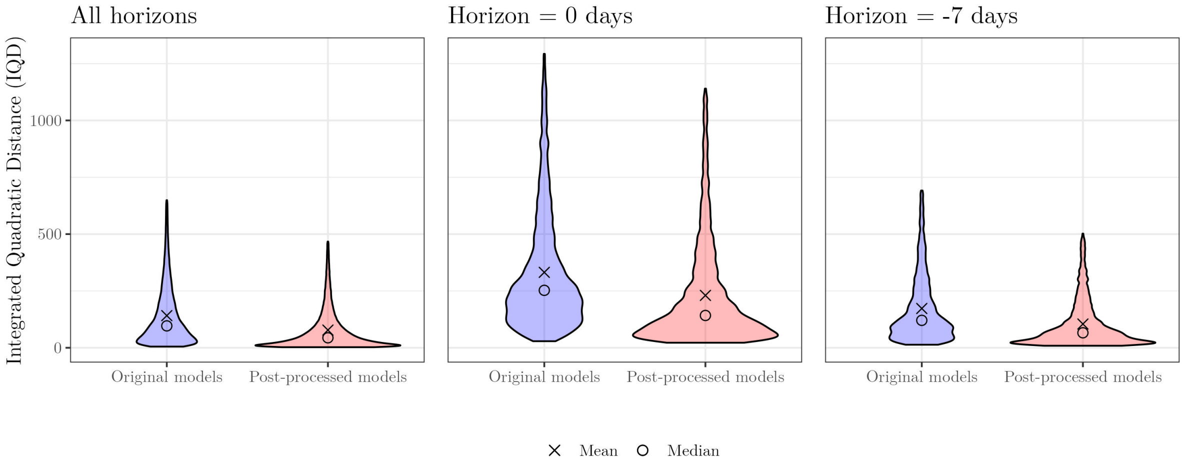


Figure SF12: Violin plots showing the average pairwise Integrated Quadratic Distance (IQD) between nowcast distributions from different models before and after post-processing (for setting `PP4`). For each pair of models, the distance is computed for each time point and horizon, and subsequently averaged. Results are shown averaged across all horizons (left), for horizon = 0 days (middle), and for horizon = $-7$ days (right). For each violin, the upper and lower 2.5% most extreme values were excluded. The symbols $\times$ and $\circ$ represent the mean and median, respectively.

### SS7 Results of DISW nowcasts

In this Section, we present the supplementary results based on the DISW ensemble methods implemented in Section 4.4.3.

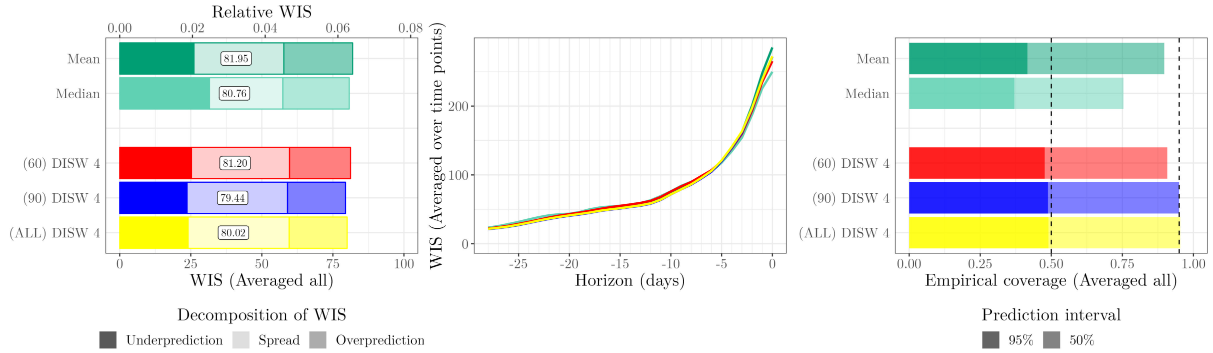


Figure SF13: Sensitivity of `DISW4` results (Figure 6) to the choice of maximum value $R$ of historical nowcast dates used in the score minimization. See caption of Figure 6 for details on plot elements.

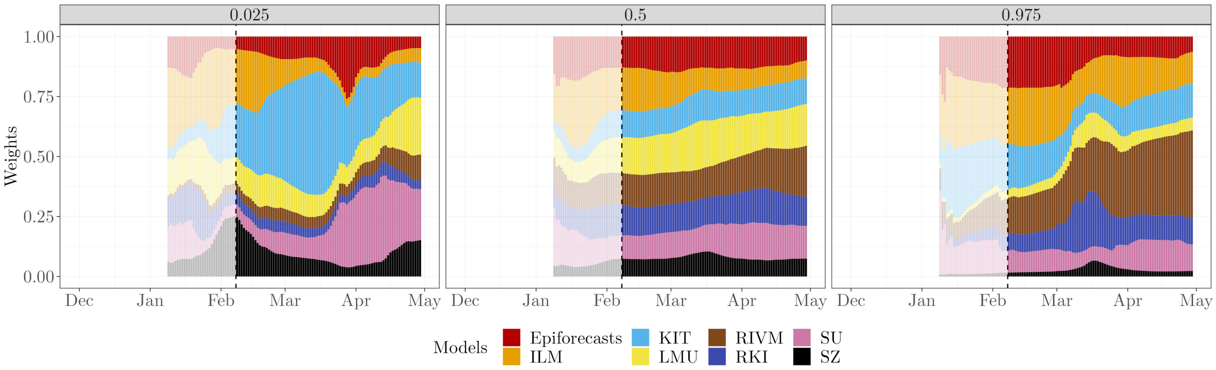


Figure SF14: Estimated weights for the 2.5$^{\text{th}}$, 50$^{\text{th}}$, and 97.5$^{\text{th}}$ percentiles based on the DISW method with weights shared across horizons while discarding incomplete observations (`DISW1`) at the national level. Similarly to Figure 5, results for the period preceding the actual evaluation period are greyed out.

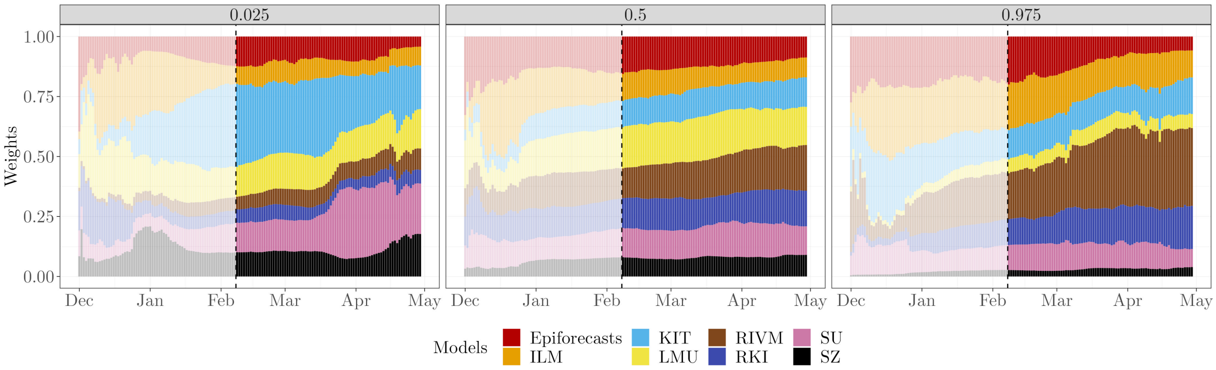


Figure SF15: Estimated weights for the 2.5$^{\text{th}}$, 50$^{\text{th}}$, and 97.5$^{\text{th}}$ percentiles based on the DISW method with weights shared across horizons with imputation with uncertainty (`DISW3`) at the national level. Similarly to Figure 5, results for the period preceding the actual evaluation period are greyed out.

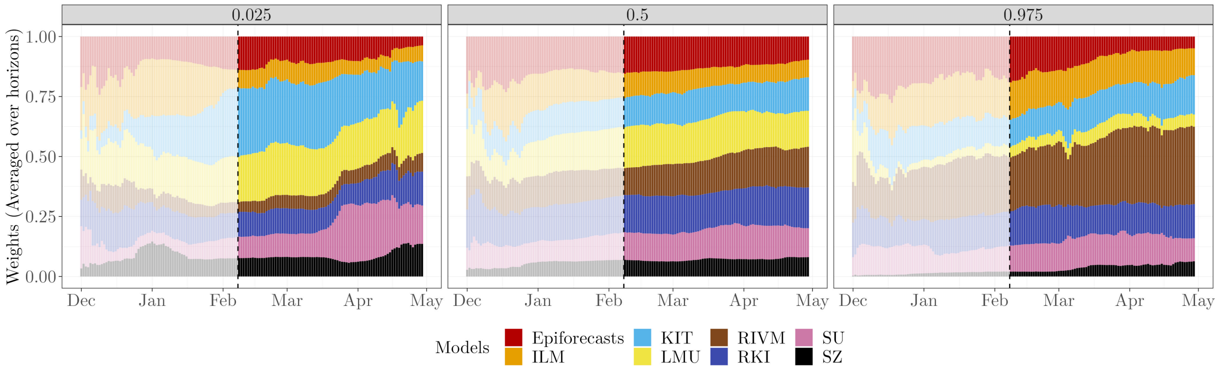


Figure SF16: Estimated weights for the 2.5$^{th}$, 50$^{th}$, and 97.5$^{th}$ percentiles based on the DISW method with weights varying over horizons with simple imputation (`DISW4`) at thenational level. Similarly to Figure 5, results for the period preceding the actual evaluation period are greyed out.

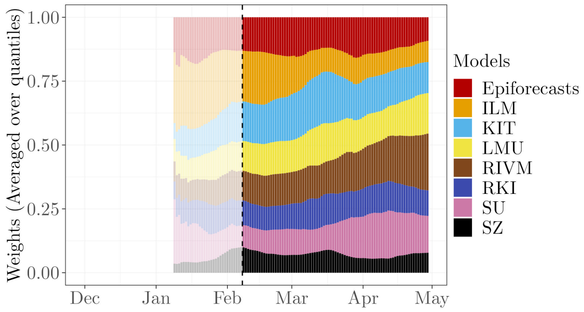


Figure SF17: Estimated weights (averaged over quantiles) based on the DISW method with weights shared across horizons while discarding incomplete observations (`DISW1`) at the national level. Similarly to Figure 5, results for the period preceding the actual evaluation period are greyed out.

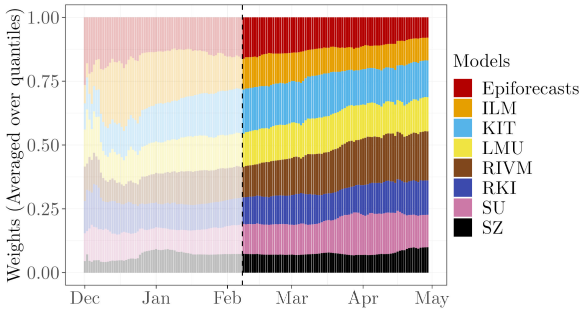


Figure SF18: Estimated weights (averaged over quantiles) based on the DISW method with weights shared across horizons with simple imputation (`DISW2`) at thenational level. Similarly to Figure 5, results for the period preceding the actual evaluation period are greyed out.

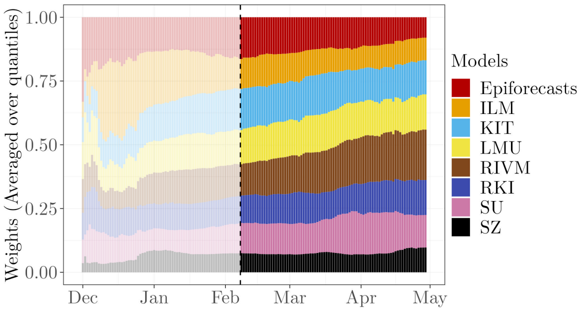


Figure SF19: Estimated weights (averaged over quantiles) based on the DISW method with weights shared across horizons with imputation with uncertainty (`DISW3`) at the national level. Similarly to Figure 5, results for the period preceding the actual evaluation period are greyed out.

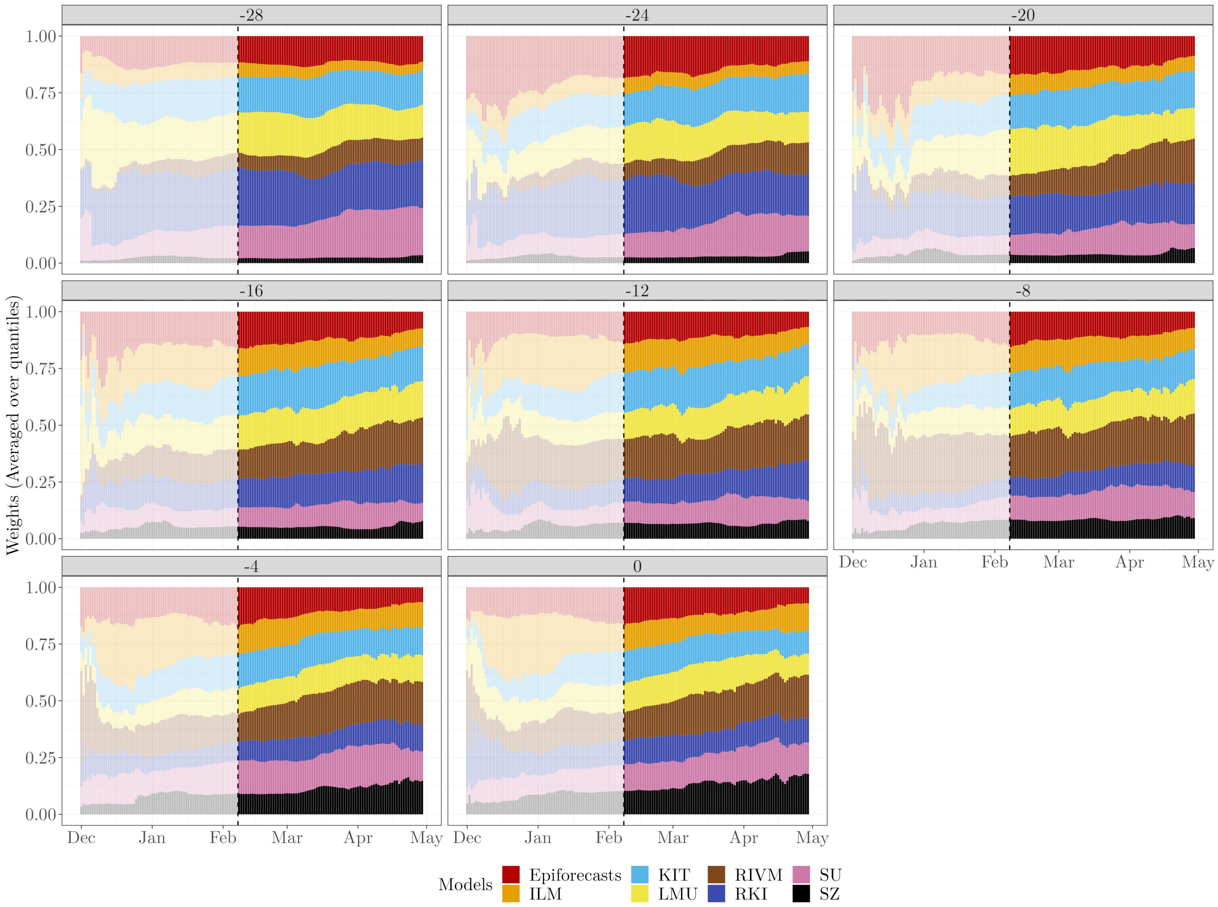


Figure SF20: Estimated weights (averaged over quantiles) with horizons of −28, −24, −20, −16, −12, −8, −4, and 0 days based on the DISW method with weights varying over horizons with simple imputation (`DISW4`) at the national level. Similarly to Figure 5, results for the period preceding the actual evaluation period are greyed out.

#### SS8 Results of AISW nowcasts

In this Section, we present the supplementary results based on the AISW ensemble methods implemented in Section 4.4.4.

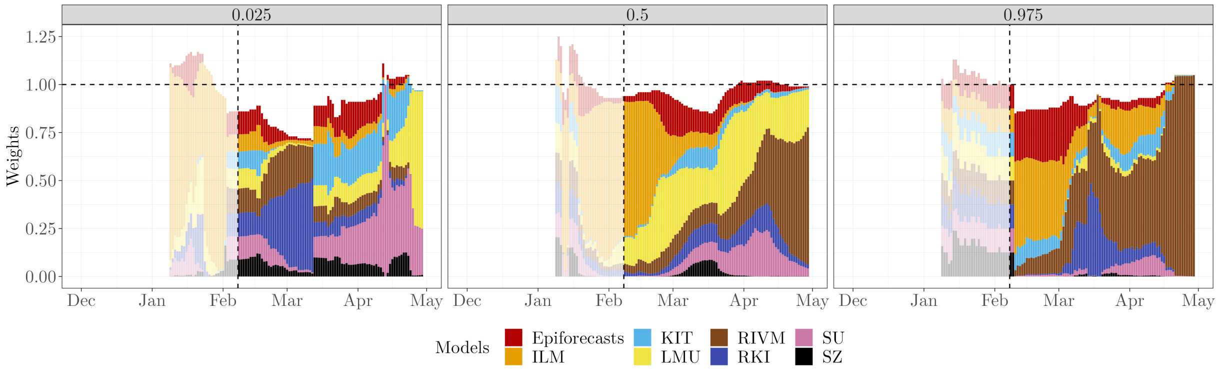


Figure SF21: Estimated weights for the 2.5th, 50th, and 97.5th percentiles based on the AISW method with weights and scaling parameter shared across horizons while discarding incomplete observations (`AISW1`) at thenational level. Similarly to Figure 5, results for the period preceding the actual evaluation period are greyed out. As a remark, due to the introduced scaling parameter $\phi^{\alpha}$, the weights are not required to sum up to 1. The horizontal dashed line represents `weight` $= 1$.

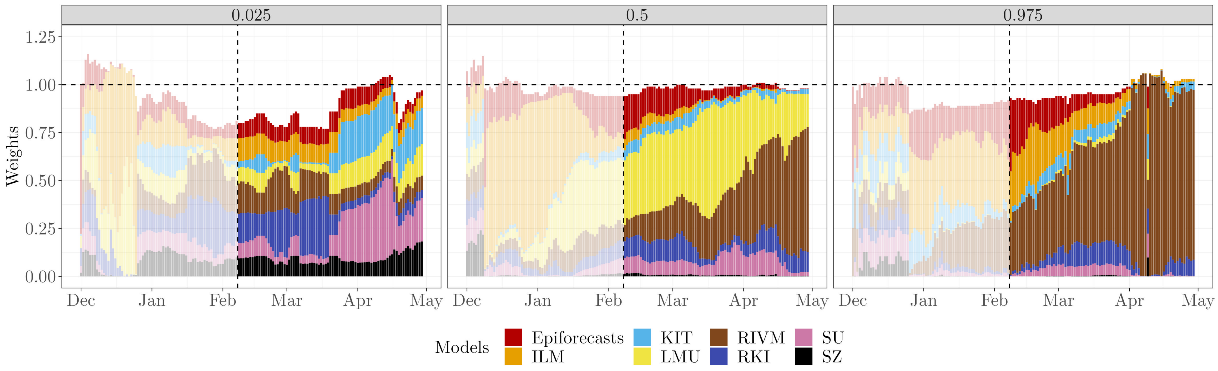


Figure SF22: Estimated weights for the 2.5th, 50th, and 97.5th percentiles based on the AISW method with weights and scaling parameter shared across horizons with imputation with uncertainty (`AISW3`) at the national level. Similarly to Figure 5, results for the period preceding the actual evaluation period are greyed out. As a remark, due to the introduced scaling parameter $\phi^{\alpha}$, the weights are not required to sum up to 1. The horizontal dashed line represents `weight` $= 1$.

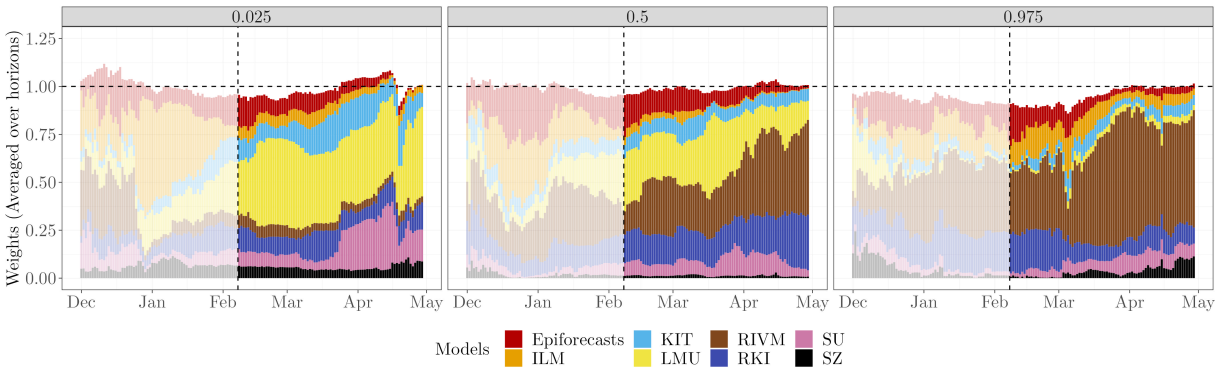


Figure SF23: Estimated weights for the 2.5$^{\text{th}}$, 50$^{\text{th}}$, and 97.5$^{\text{th}}$ percentiles based on the AISW method with weights and scaling parameter varying over horizons with simple imputation (`AISW4`) at the national level. Similarly to Figure 5, results for the period preceding the actual evaluation period are greyed out. As a remark, due to the introduced scaling parameter $\phi^{\alpha,t^\star-t}$, the weights are not required to sum up to 1. The horizontal dashed line represents `weight` $= 1$.

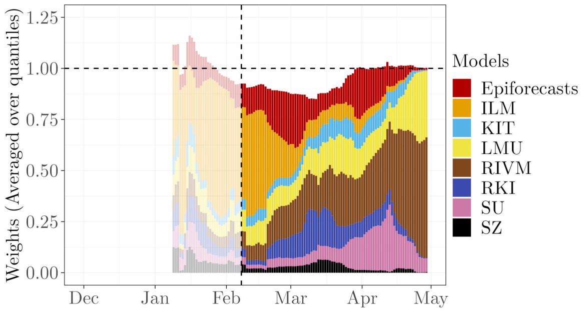


Figure SF24: Estimated weights (averaged over quantiles) based on the AISW method with weights shared across horizons while discarding incomplete observations (`AISW1`) at the national level. Similarly to Figure 5, results for the period preceding the actual evaluation period are greyed out. As a remark, due to the introduced scaling parameter $\phi^{\alpha}$, the weights are not required to sum up to 1. The horizontal dashed line represents `weight` $= 1$.

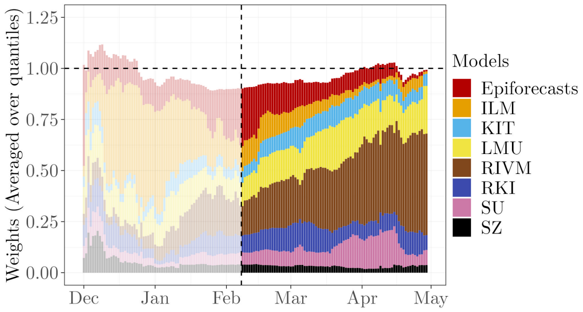


Figure SF25: Estimated weights (averaged over quantiles) based on the AISW method with weights shared across horizons with simple imputation (`AISW2`) at the national level. Similarly to Figure 5, results for the period preceding the actual evaluation period are greyed out. As a remark, due to the introduced scaling parameter $\phi^{\alpha}$, the weights are not required to sum up to 1. The horizontal dashed line represents `weight` $= 1$.

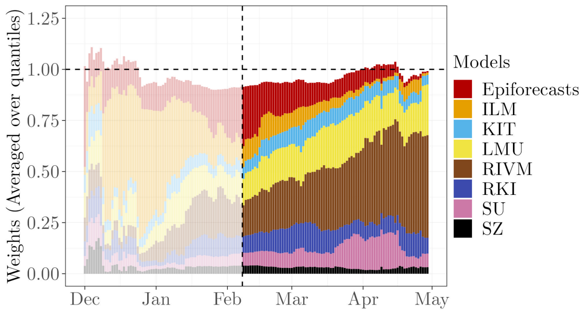


Figure SF26: Estimated weights (averaged over quantiles) based on the AISW method with weights shared across horizons with imputation with uncertainty" (`AISW3`) at the national level. Similarly to Figure 5, results for the period preceding the actual evaluation period are greyed out. As a remark, due to the introduced scaling parameter $\phi^{\alpha}$, the weights are not required to sum up to 1. The horizontal dashed line represents `weight` $= 1$.

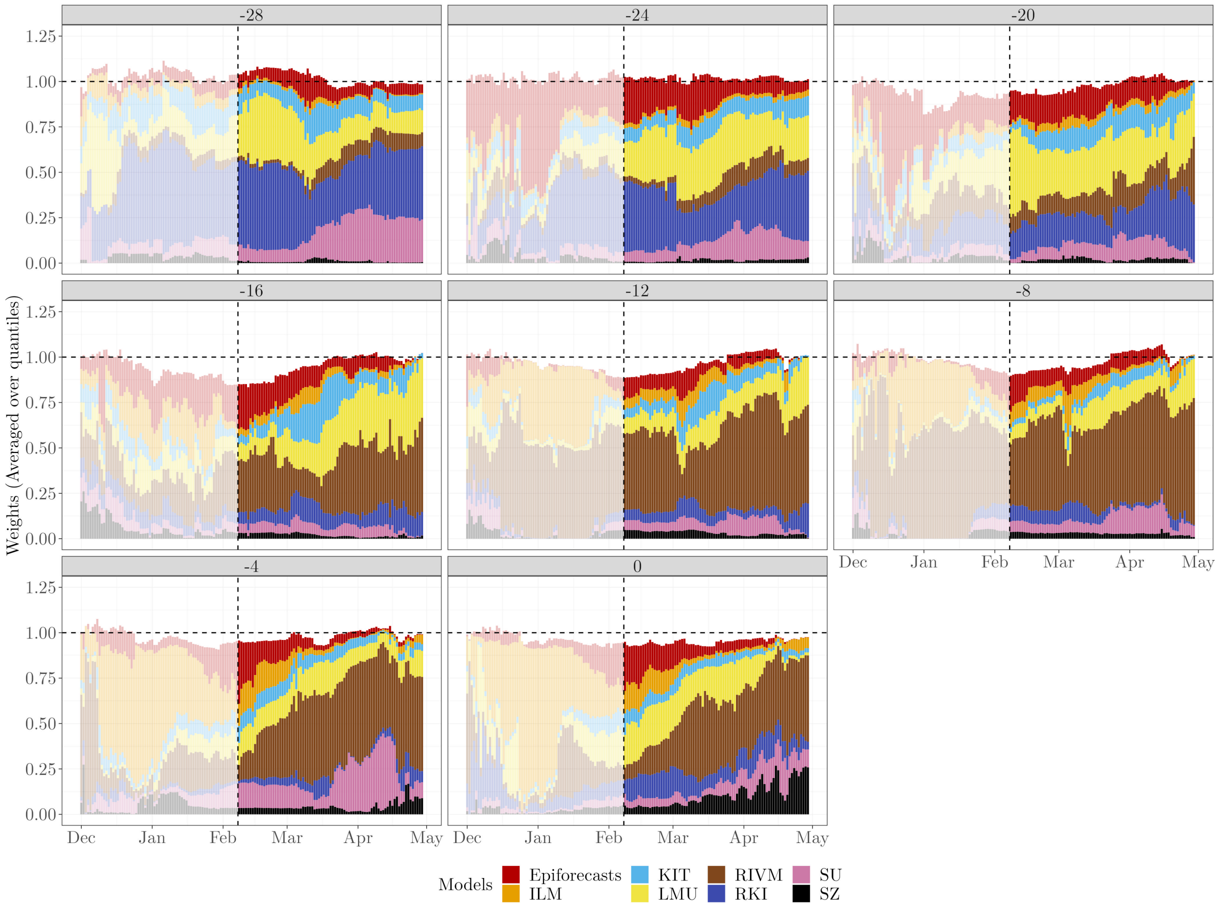


Figure SF27: Estimated weights (averaged over quantiles) with horizons of $-28$, $-24$, $-20$, $-16$, $-12$, $-8$, $-4$, and 0 days based on the AISW method with weights varying over horizons with simple imputation (`AISW4`) at the national level. Similarly to Figure 5, results for the period preceding the actual evaluation period are greyed out. As a remark, due to the introduced scaling parameter $\phi^{\alpha,t^\star-t}$, the weights are not required to sum up to 1. The horizontal dashed line represents `weight` $= 1$.

#### SS9 Results of Select-$n$

In this Section, we present the supplementary results based on the select-$n$ approach implemented in Section 4.4.5.

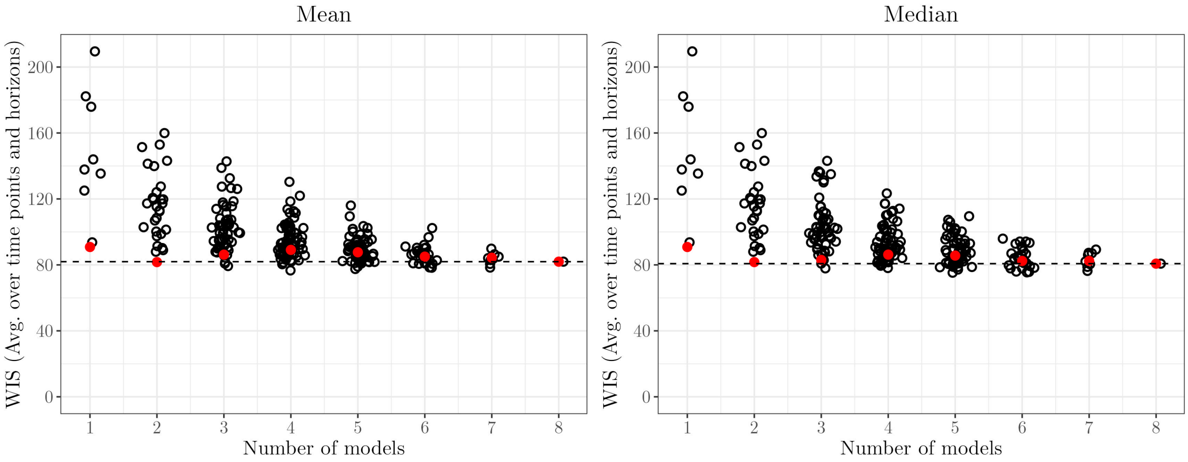


Figure SF28: Results corresponding to Figure 11, but with model selection done jointly for all horizons (`Select-n-Mean1` and `Select-n-Median1`) rather than separately per horizon.
